# Mendelian randomization analyses uncover causal relationships between brain structural connectome and risk of psychiatric disorders

**DOI:** 10.1101/2025.02.20.25322606

**Authors:** Kanwei Xiao, Xinle Chang, Chenfei Ye, Zhiguo Zhang, Ting Ma, Jingyong Su

**Affiliations:** School of Computer Science and Technology, Harbin Institute of Technology (Shenzhen), Shenzhen, China; School of Biomedical Engineering, Harbin Institute of Technology (Shenzhen), Shenzhen, China; Pengcheng Laboratory, Shenzhen, China

## Abstract

Growing evidence suggests abnormalities of brain structural connectome in psychiatric disorders, but the causal relationships remain underexplored. We conducted bidirectional two-sample Mendelian randomization (MR) analyses to investigate the causal links between 206 white-matter connectivity phenotypes (n = 26,333, UK Biobank) and 13 major psychiatric disorders (n = 14,307 to 1,222,882). Forward MR analyses identified causal effects of genetically predicted five white-matter structural connectivity phenotypes on six psychiatric disorders, with associations being significant or suggestive. For instance, structural connectivity between the left-hemisphere frontoparietal control network and right-hemisphere default mode network was significantly negatively associated with autism spectrum disorder risk, while increased structural connectivity between the right-hemisphere frontoparietal control network and hippocampus was significantly linked to decreased anorexia nervosa and cannabis use disorder risk. Reverse MR analyses revealed significantly or suggestively causal relationships between the risk of two psychiatric disorders and four different white-matter structural connectivity phenotypes. For example, the susceptibility of anorexia nervosa was found to be significantly negatively associated with structural connectivity between the left-hemisphere visual network and pallidum. These findings offer new insights into the etiology of psychiatric disorders and highlight potential biomarkers for early detection and prevention at the brain structural connectome level.

## Introduction

Psychiatric disorders are a group of mental illnesses that manifest as dysfunctions in emotional regulation, cognition or behavior^1^. Due to their high prevalence, mortality and morbidity risk, psychiatric disorders represent a global public health threat that imposes economic burdens worldwide^2,3^. However, our understanding of the etiology of psychiatric disorders remains limited, which impedes the development of effective therapeutic strategies.

Alterations within the brain structural connectome have been extensively reported to be induced by psychiatric disorders. For example, reductions in structural connectivity within a network involving the frontal, striatal, and cerebellar regions have been observed in attention-deficit/hyperactivity disorder (ADHD) patients^4^. Compared to healthy controls, individuals with anorexia nervosa (AN) exhibit decreased connectivity within subcortical networks and enhanced connectivity between frontal cortical regions^5^. Generalized anxiety disorder is reported to be associated with disruptions in a structural sub-network primarily involving the frontal-subcortical circuits, which may serve as a potential neuroimaging biomarker for diagnosis^6^. Investigations have revealed that high-functioning autism spectrum disorder (ASD) patients have less white-matter density in the anterior part of the corpus callosum than typically developing peers^7^. In patients with bipolar disorder (BIP), enhanced white-matter connectivity is observed between the left subgenual cingulate and left amygdalo-hippocampal complex (including the amygdala, hippocampus, and associated regions) compared to healthy individuals^8^. Reduced white-matter connectivity in the default mode network and the frontal-thalamus-caudate regions has been reported in patients with depression^9^. Individuals with obsessive-compulsive disorder (OCD) exhibit reduced structural connectivity in a network primarily involving orbitofrontal, striatal, insula and temporo-limbic regions, which are implicated in the disorder’s pathophysiology^10^. Post-traumatic stress disorder (PTSD) is reported to be associated with altered structural connectivity, including potential nodal centrality decreases in the medial orbital part of the superior frontal gyrus and increases in the salience network^11^. In schizophrenia (SCZ), decreased connectivity in frontal and temporal regions is observed, along with a diminished central role of frontal hubs in the brain network^12^. Tourette syndrome (TS) patients exhibit elevated structural connectivity between the striatum, thalamus and multiple brain regions, including the motor and sensory cortices, paracentral lobule, supplementary motor area, and parietal cortices^13^. Alcohol use disorder (AUD) patients are observed to have lower white-matter integrity in the cerebellum and right insula, and increased white-matter connectivity in the default mode network^14^. Cannabis use disorder (CUD) is associated with disrupted structural connectivity in the fornix, corpus callosum and commissural fibers^15^. Patients with heroin use disorder, a subtype of opioid use disorder (OUD), exhibit increased structural connectivity in the paralimbic, orbitofrontal, prefrontal, and temporal regions^16^. The observed findings delineate the presence of dysconnectivity within the brain’s structural connectivity patterns in diverse psychiatric disorders. Nonetheless, the causal relationships between these structural networks and the psychiatric disorders remains largely unexplored.

The gold standard for studying causal relationships is randomized controlled trials (RCTs). However, due to limitations such as cost and ethical concerns, RCTs are not always feasible. With the increasing availability of large-scale genome-wide association studies (GWAS), MR has gained prominence as a valuable alternative to RCTs^17^. MR utilizes genetic variations (typically single nucleotide polymorphisms, or SNPs) associated with an exposure as instrumental variables (IVs) to evaluate the causal effect of exposure on the outcome^18^. Compared to conventional observational studies, MR analysis offers a methodological advantage by substantially reducing biases arising from confounding factors and eliminating reverse causation. This enhanced validity stems from the fundamental biological principle that genetic alleles undergo random segregation during meiosis, and genetic variants are determined prior to both exposure and outcome variables^17,18^. Previous MR studies have primarily focused on examining the relationships between white-matter tract microstructure and psychiatric disorders^19,20^. However, these investigations are limited by the fact that white-matter fiber tracts only provide indirect measures of anatomical connectivity without explicitly characterizing the complex interregional relationships within the brain. In contrast, the structural connectome offers a comprehensive representation of whole-brain connectivity, enabling global detection of brain network reorganization^21^. Therefore, investigating the causal links between inter-regional brain connectivity and psychiatric disorders shows a remarkable advancement in understanding the neurobiological basis of mental illnesses.

In this study, we conducted a bidirectional two-sample MR analyses to explore the potential causal links between the brain structural connectome and 13 major psychiatric disorders. The brain structural connectome encompasses 206 white-matter connectivity metrics, quantifying the density of white-matter tracts that interconnect cortical hemispheres, cortical networks, and subcortical regions, either internally or across these modules. Our findings might provide novel insights into the neuropathological mechanisms underlying major psychiatric disorders through the lens of brain structural connectomics. These discoveries have significant translational implications, potentially informing the development of: (1) early diagnostic biomarkers, (2) targeted intervention strategies, and (3) personalized treatment approaches based on individual connectome profiles.

## Results

### Overview of this study

Our study design is briefly illustrated in Fig.1. To systematically investigate potential causal relationships between brain structural connectome organization and psychiatric disorders, we implemented a comprehensive bidirectional two-sample MR framework. This analyses leveraged the largest GWAS dataset currently available, comprising 206 distinct white-matter structural connectivity phenotypes that capture interregional connectivity patterns across multiple brain networks and anatomical divisions^21^. These phenotypes include: (1) 3 hemisphere-level connectivity measures (intra- and inter-hemispheric); (2) 105 network-level measures, with 14 intra-network and 91 inter-network connectivity across seven bilateral networks^22^; and (3) 98 network-to-subcortical measures between cortical networks and seven subcortical structures (see Supplementary Table 1 for details). Furthermore, we incorporated large-scale GWAS summary statistics for 13 psychiatric disorders, selected to minimize sample overlap and restricted to individuals of European ancestry (see Table 1 and Supplementary Table 2 for details). Although there remains a maximum possible sample overlap of approximately 5.29% for AN^23^ and 2.15% for PTSD^24^, these minimal overlaps are deemed insufficient to significantly bias the study results^19,25,26^. To satisfy the IV assumptions in MR analyses^27^, we conducted a rigorous IV selection and removed outliers. All significant exposure-outcome pairs reported had F statistics greater than 30 (see Supplementary Tables 3 and 4 for details), indicating robust IVs. Forward MR analyses identified six putative causal associations, while reverse MR analyses revealed four putative causal associations. Considering multiple comparisons, the Bonferroni-corrected significance threshold was set at 1.2316 × 10^−4^ (0.05/206/2, 206 denotes the number of white-matter structural connectivity phenotypes, 2 represents bidirectional MR analyses). Meanwhile, results slightly below the Bonferroni threshold may still be of suggestive value, and a nominal significance threshold was set at 1 × 10^−3^. A series of sensitivity analyses confirmed the robustness of our results. Lastly, we employed birth length^28^ in our analyses. Our findings revealed no significant associations with white-matter structural connectivity after Bonferroni correction, nor any nominally significant associations with the 13 psychiatric disorders. These results further validate the reliability of our causal inferences.

**Fig. 1.**
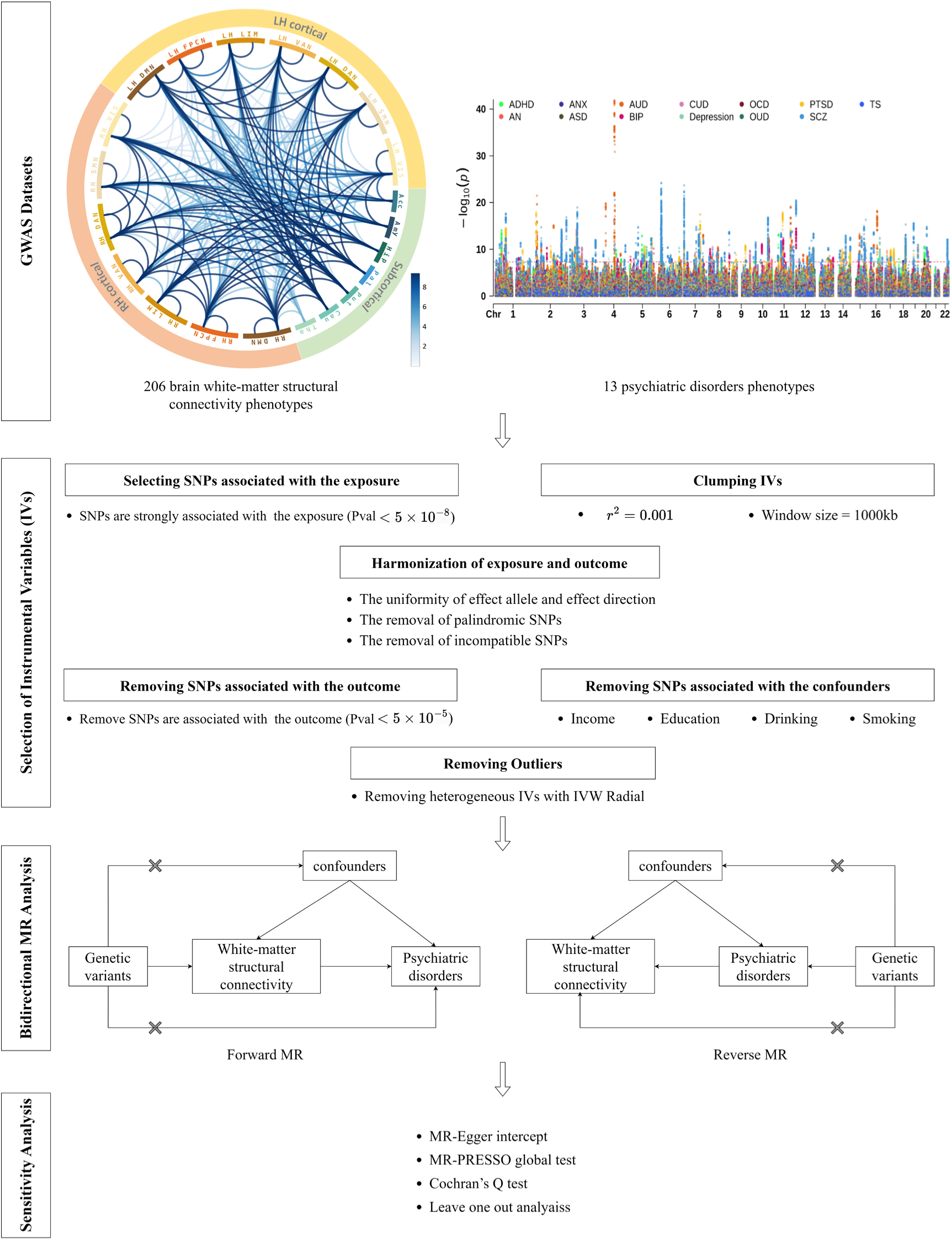
Study flowchart for bidirectional MR analyses between brain structural connectome and psychiatric disorders. A total of 206 white-matter structural connectivity phenotypes and 13 major psychiatric disorders were included for causality inference. IVs were selected based on their strong association with the exposure and independence after clumping to remove linkage disequilibrium, excluding any SNPs associated with confounders or the outcome. Outliers showing significant heterogeneity were further discarded. Bidirectional MR analyses were performed to investigate causal links between white-matter structural connectivity phenotypes and psychiatric disorders. To validate the reliability and consistency of the MR findings, a series of sensitivity analyses were conducted. The top left chord plot visualizes the average values of the brain structural connectivity measures across GWAS participants. The top right Manhattan plot visualizes the GWAS summary data of 13 psychiatric disorders. Abbreviations: LH, left hemisphere; RH, right hemisphere; VIS, visual network; SMN, somatomotor network; DAN, dorsal attention network; VAN, ventral attention network; LIM, limbic network; FPCN, frontoparietal control network; DMN, default mode network; Tha, thalamus; Cau, caudate; Put, putamen; Pal, pallidum; Hip, hippocampus; Amy, amygdala; Acc, accumbens.

**Table 1.**
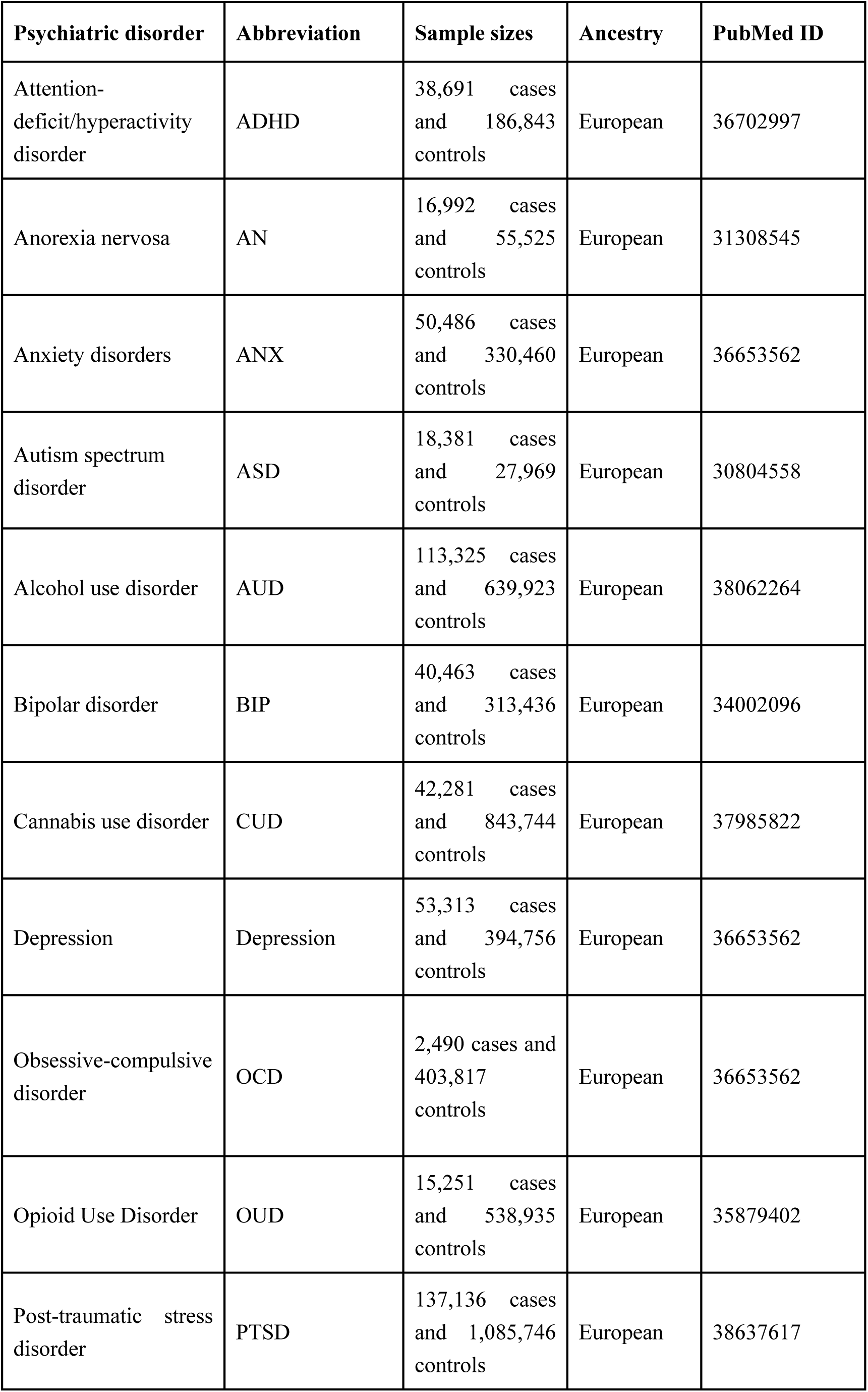
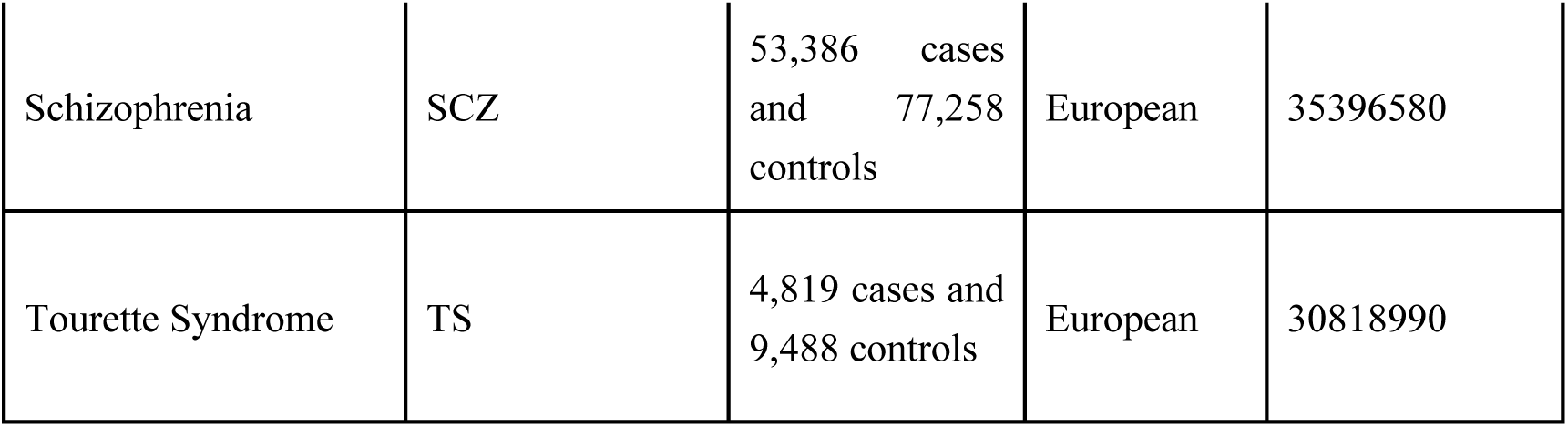
Overview of the GWAS summary data for psychiatric disorders.

### Forward MR results of brain structural connectome on psychiatric disorders

We identified six putative causal links between white-matter structural connectivity phenotypes and risk of psychiatric disorders in the forward MR analyses, as shown in Fig. 2 and Supplementary Table 7. The IVW estimates suggest that genetically determined white-matter structural connectivity between the left-hemisphere dorsal attention network (DAN) and right-hemisphere somatomotor network (SMN) was nominally negatively associated with the risk of ADHD (IVW OR = 0.64, 95% CI: 0.51 to 0.81, *P* = 1.88 × 10^−4^). And genetically determined white-matter structural connectivity between the right-hemisphere frontoparietal control network (FPCN) and hippocampus was significantly negatively associated with the risk of AN (IVW OR = 0.50, 95% CI: 0.37 to 0.68, *P* = 1.11 × 10^−5^). Nominally positive causal association was also observed between the left-hemisphere FPCN to the right-hemisphere FPCN white-matter structural connectivity and the risk of anxiety disorders (IVW OR = 1.29, 95% CI: 1.12 to 1.49, *P* = 6.35 × 10^−4^). Additionally, white-matter structural connectivity between the left-hemisphere FPCN and right-hemisphere default mode network (DMN) was found to be significantly negatively associated with the risk of ASD (IVW OR = 0.55, 95% CI: 0.41 to 0.73, *P* = 3.59 × 10^−5^). Moreover, significantly negative association was observed between white-matter structural connectivity linking the right-hemisphere FPCN with hippocampus and the risk of CUD (IVW OR = 0.47, 95% CI: 0.35 to 0.64, *P* = 9.66 × 10^−7^). Furthermore, genetically determined cross-hemisphere white-matter structural connectivity was nominally negatively associated with the risk of SCZ (IVW OR = 0.61, 95% CI: 0.46 to 0.82, *P* = 7.61 × 10^−4^). The statistical power of significant results in forward MR analyses ranged from 98.6% to 100% (see Supplementary Table 5 for details), which demonstrated the reliability of our findings.

**Fig. 2.**
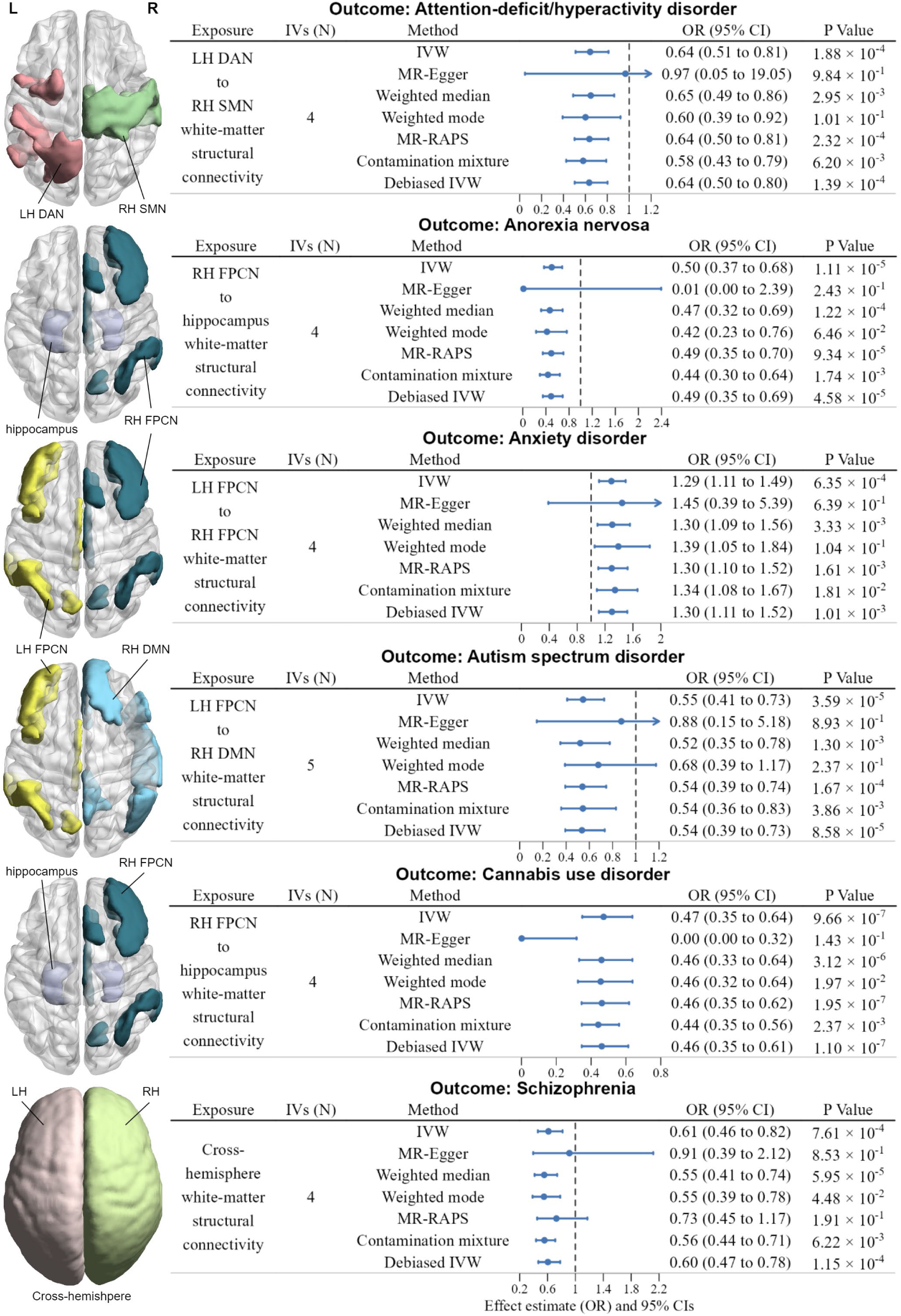
Significant results of forward MR analyses. **Left**: Axis view (dorsal side) of related functional networks and subcortical structures. L, left; R, right. **Right**: forest plot illustrates the Bonferroni-corrected significant (*P* < 1.2316 × 10^−4^) and nominally significant (*P* < 1 × 10^−3^) IVW results for the causal effects of white-matter structural connectivity on psychiatric disorders, along with the results from six additional methods: MR-Egger, Weighted median, Weighted mode, MR-RAPS, Contamination mixture, Debiased IVW. Arrows indicate the extension of the maximum interval on the x-axis. OR refers to odds ratio, and the error bars represent the 95% CIs (confidence intervals). P values were from each MR analyses method, and all statistical tests were two-sided.

### Reverse MR results of psychiatric disorders on brain structural connectome

We also identified four putative causal relationships between psychiatric disorders and white-matter structural connectivity phenotypes in the reverse MR analyses, as shown in Fig. 3 and Supplementary Table 8. Significantly negative causal effect of the susceptibility to AN on white-matter structural connectivity between the left-hemisphere visual network and pallidum was observed (IVW beta = −0.19, 95% CI: −0.26 to −0.11, *P* = 7.76 × 10^−7^). Moreover, genetically predicted increased susceptibility to SCZ was nominally associated with the higher white-matter structural connectivity between the left-hemisphere DMN and putamen (IVW beta = 0.04, 95% CI: 0.02 to 0.06, *P* = 2.36 × 10^−4^), nominally associated with the higher white-matter structural connectivity between the left-hemisphere visual network and putamen (IVW beta = 0.04, 95% CI: 0.02 to 0.07, *P* = 2.84 × 10^−4^), and nominally associated with the higher white-matter structural connectivity between the right-hemisphere limbic network and amygdala (IVW beta = 0.03, 95% CI: 0.01 to 0.05, *P* = 6.04 × 10^−4^). The statistical power of significant results in reverse MR analyses ranged from 24.5% to 43.6%, as detailed in Supplementary Table 6. This relatively low power is primarily attributed to the small effect sizes of psychiatric disorders on white-matter structural connectivity. Specifically, the beta values were close to zero, indicating weak causal associations between psychiatric disorders and white-matter connectivity.

**Fig. 3.**
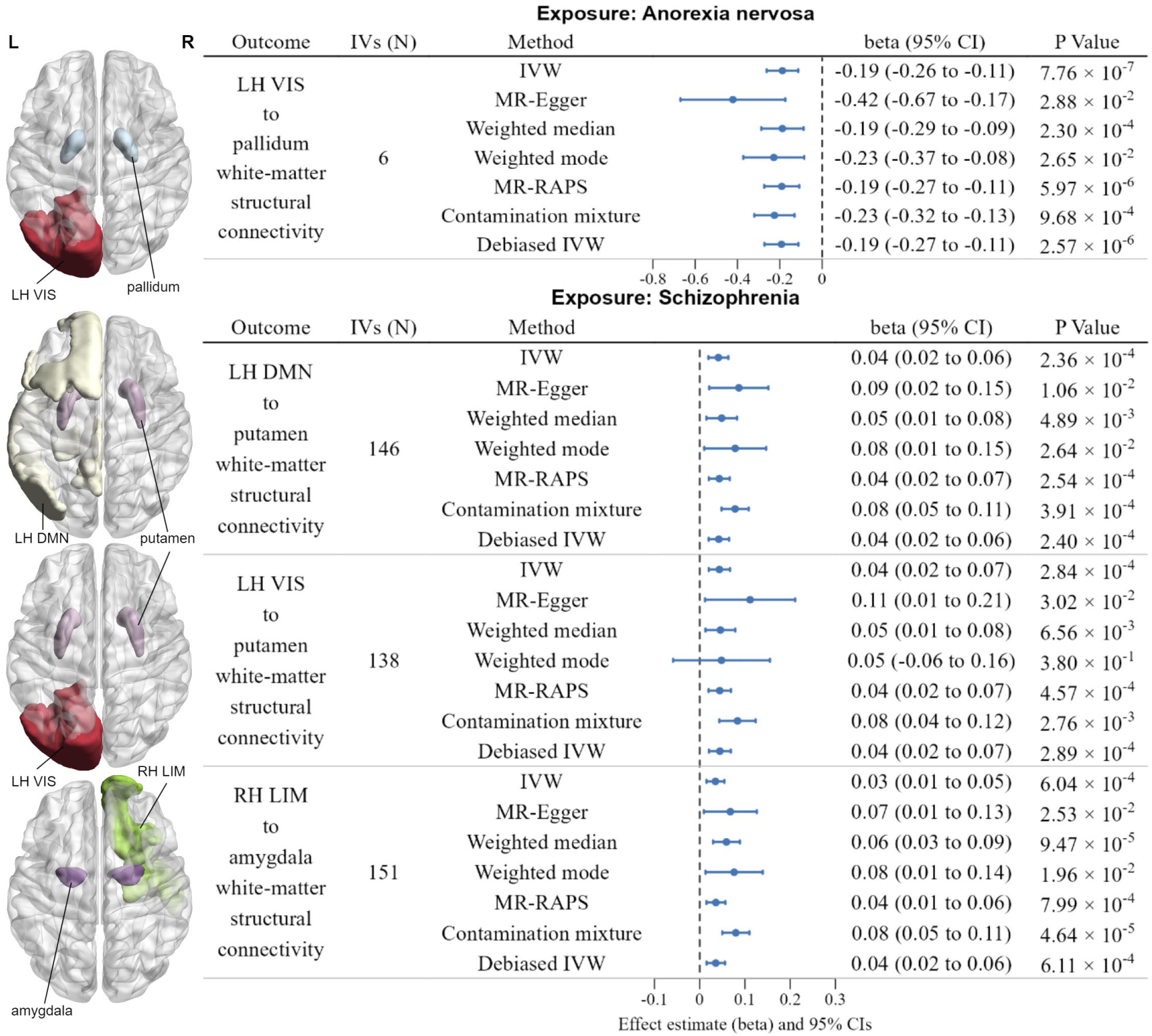
Significant results of reverse MR analyses. **Left**: Axis view (dorsal side) of related functional networks and subcortical structures. **Right**: forest plot illustrates the Bonferroni-corrected significant (*P* < 1.2316 × 10^−4^) and nominally significant (*P* < 1 × 10^−3^) IVW results for the causal effects of psychiatric disorders on white-matter structural connectivity, along with the results from six additional methods: MR-Egger, Weighted median, Weighted mode, MR-RAPS, Contamination mixture, Debiased IVW. The error bars represent the 95% CIs (confidence intervals). P values were from each MR analyses method, and all statistical tests were two-sided.

### Sensitivity analyses

Six different MR methods—MR-Egger, weighted median, weighted mode, MR-robust adjusted profile scores (MR-RAPS), contamination mixture, and debiased inverse variance weighted (Debiased IVW)—yielded consistent causal effect directions with IVW, further supporting the robustness of the causal inference (see Supplementary Tables 7 and 8, and Supplementary Figures 1 to 10). Not all MR methods produced statistically significant results, likely due to their lower statistical power compared to the IVW method^29,30^.

Additional sensitivity analyses validated the reliability of the results. MR-Egger regression and the MR pleiotropy residual sum and outlier (MR-PRESSO) global test detected no evidence of horizontal pleiotropy or outliers in the reported significant exposure-outcome pairs in bidirectional MR analyses. Moreover, Cochran’s Q test revealed no significant heterogeneity across different IVs. The leave-one-out analyses indicated that no single IV disproportionately influenced the results, and the causal effects remained consistent when each IV was excluded. The sensitivity analyses results are available in Supplementary Tables 5 and 6, with the exception of the leave-one-out analyses, which are presented in Supplementary Figures 1 to 10.

## Discussion

This MR study provides novel insights into the bidirectional causal relationships between brain structural connectome organization and psychiatric disorders. Leveraging publicly available GWAS summary statistics, we conducted systematic bidirectional two-sample MR analyses encompassing 206 white-matter structural connectivity phenotypes and 13 major psychiatric disorders. Our investigation revealed significant causal associations, including: (1) five specific white-matter connectivity patterns showing causal effects on six distinct psychiatric disorders, and (2) two psychiatric disorders demonstrating causal influences on four different white-matter structural connectivity profiles. To ensure the robustness of our findings, we implemented a rigorous series of sensitivity analyses, including consistency checks of causal directions across multiple MR methods, pleiotropy detection, heterogeneity testing, and leave-one-out analyses, all of which reliably supported the validity of our primary results.

ADHD, marked by difficulties in sustaining attention or by impulsive and hyperactive behaviors, is increasingly conceptualized as a brain dysconnectivity disorder. In our forward MR analyses, we identified that decreased bilateral brain structural connectivity between the DAN and the SMN was positively associated with the risk of ADHD. The DAN, comprising key regions including the dorsal and lateral prefrontal cortex, superior parietal lobule, and intraparietal sulcus, has been consistently implicated in ADHD pathophysiology, with disrupted microstructural properties serving as established connectomic signatures of the disorder^31^. Meanwhile, microstructural characteristics in the SMN have been reported to play a crucial role in attention and impulsivity in ADHD population^32^. In addition to WM structural connectivity findings, functional interconnections between DAN and SMN were reported associated with impulsive behaviors in children with ADHD^33^. Notably, prior meta-analytic evidence suggests a developmental shift in the neural correlates of ADHD, with childhood manifestations primarily characterized by SMN hypoactivation that transitions to predominant DAN hypoactivation in adulthood^34^. Further longitudinal clinical studies focusing on these two brain networks may help to confirm this developmental variation and elucidate its implications for ADHD patients. Altogether, our MR-based evidence indicates that altered structural connectivity between the DAN and the SMN may play a vital role in ADHD.

Our bidirectional Mendelian randomization analyses revealed a complex neurobiological interplay underlying AN, characterized by energy restriction behaviors and distorted body perception. The forward MR demonstrated that decreased connectivity between the FPCN and hippocampus confers heightened AN risk. The hippocampus is thought to be involved in the energy intake and weight-regulation processes^35^, probably explaining alterations of eating behaviors in AN patients^36,37^.

Congruently, anatomical MRI studies also reported volume reduction in the hippocampus^38–40^ and disrupted microstructures of white matter fiber (e.g., fornix, cingulum) linking to the hippocampus in AN patients^37,41–43^. Of note, a recent study has suggested that brain activity within FPCN may function as a biomarker to predict treatment response in individuals with AN^44^. Cortical thinning of these regions observed in AN populations^38^, combining with our findings regarding structural connectivity, indicates that the functional aberrations of FPCN in AN may come from the underlying structural damages. The reverse MR analyses further identified AN-induced hypoconnectivity between the pallidum and the visual network. Although no direct evidence has been reported linking the pallidum-visual connection to AN, decreased structural connectivity within this neural pathway has been primarily associated with obsessive-compulsive personality disorder, which is often comorbid with AN^45^. Based on the neural basis of the ventral pallidum in regulating food motivation and reward^46,47^, as well as the role of the ventral visual network in subserving the visual perception of the body^48,49^, we can infer that the compulsive disorder-related behavior may be induced after the onset of AN through the disruption of the pallidum-visual connection. In sum, our results revealed a complex pattern of connectomic signatures in AN, offering novel insights into the pathophysiology of AN.

Anxiety disorders (ANX), characterized by maladaptive fear responses and stimulus avoidance patterns, demonstrate associations with disruptions in brain network connectivity. Our forward MR analyses identified increased structural connectivity within bilateral FPCN as a causal factor for higher ANX risk. Neuroimaging evidence implicates the central role of prefrontal cortex in both the pathophysiology^50^ and prediction^51^ of ANX. In social anxiety disorder, the genu of the corpus callosum, which bridges bilateral prefrontal cortical regions, exhibits elevated white-matter density when seeded in the right medial prefrontal cortex^52^. Additionally, the dorsal anterior cingulate cortex, anatomically anterior to the genu and functionally connected with the FPCN^53^, plays a key role in attentional control and emotional regulation^52^. Abnormally heightened thickness in FPCN areas^54,55^ and prefrontal-limbic hyperconnectivity patterns^51,56^ were observed in anxiety disorders patients, although these findings are not entirely consistent across studies^6,57–59^. Our results contribute to addressing this heterogeneity and offer a potential neural connectivity basis for ANX, particularly in relation to inter-hemispheric hyperconnectivity within the FPCN. The heightened structural connectivity, as indexed by streamline density^21^, might indicate impaired axonal pruning during critical developmental periods^60^ or aberrant myelination triggered by neurobiological dysfunction^61^. This could reflect inefficiency in FPCN connectivity during anxiety-related cognitive process or maladaptive adaptation to underlying neurobiological deficits.

ASD is characterized by social communication deficits and restricted/repetitive behaviors, with emerging evidence implicating brain network reorganization in its pathophysiology. Our forward MR analyses identified a potential causal relationship between diminished structural connectivity linking the left FPCN with the right DMN and increased ASD susceptibility. This finding aligns with longitudinal neuroimaging evidence demonstrating divergent developmental trajectories of FPCN-DMN connectivity: neurotypical individuals demonstrate normative age-associated strengthening of these inter-network connections, while high-functioning ASD patients experience a progressive decline of FPCN-DMN interconnection^62^. Moreover, volumetric variations within FPCN and DMN regions correlate with intelligence quotient development in ASD, suggesting network-specific neuroanatomical biomarkers^63^. Complementary literature reveals that ASD is linked to abnormalities in white-matter integrity within tracts connecting regions associated with executive control functions (e.g., the FPCN) and socio-emotional processing (e.g., the DMN)^64^. Specifically, diffusion magnetic resonance imaging studies have highlighted disruptions in several critical white-matter pathways, including the cingulum bundle^65^, corpus callosum, uncinate fasciculus, and superior longitudinal fasciculus^66^. These convergent brain structural reorganization patterns provide mechanistic context for our MR-derived hypothesis of bilateral FPCN-DMN hypoconnectivity contributing to ASD pathogenesis.

In our forward MR analyses, reduced white-matter connectivity between the FPCN and hippocampus was found to be causally associated with an elevated risk of CUD. Consistent with this, previous studies have reported structural impairments in both the frontoparietal^67,68^ and hippocampus^69,70^ areas among cannabis users. Researchers further postulates that the shift from voluntary to habitual drug consumption may stem from disruptions in brain regions governing executive control and behavioral inhibition^71^. The FPCN, which plays a crucial role in decision-making and inhibitory regulation^72,73^, might be among these affected regions. Interestingly, our study discovered that the structural connectivity between right-hemisphere FPCN and hippocampus converges as a negative causal factor influencing the risks of both AN and CUD. Consistent with our MR findings that implicate shared neural pathways, epidemiological studies have reported a 14% prevalence of cannabis use and a 6% prevalence of CUD in AN patients^74^, suggesting a possible comorbid relationship between the two conditions. Genetic analyses further suggest that individuals with a genetic predisposition to AN may exhibit a similar vulnerability to developing CUD^75^. Emerging evidence has proposed cannabis use as a potential therapeutic intervention for AN symptoms, particularly in addressing weight restoration and associated physiological complications^76,77^. However, the potential adverse effects of cannabis use in this context warrant careful consideration, including the risk of precipitating binge episodes and subsequent compensatory behaviors, especially when individuals experience post-consumption guilt regarding their eating patterns^78^. An alternative hypothesis posits that symptoms of cannabinoid hyperemesis syndrome might be misdiagnosed as compensatory behaviors in individuals with binge-eating/purging subtype AN, due to their overlapping clinical presentations^79^. These findings underscore the need for future clinical research to resolve existing inconsistencies and to develop personalized cannabis dosing strategies that maximize therapeutic efficacy while minimizing potential adverse effects.

The current study revealed widespread disturbances in both cortico-cortical and cortico-subcortical structural connectivity in patients with SCZ. In the forward MR, we found that inter-hemispheric hypoconnectivity was associated with the risk of SCZ, echoing the well-recognized lateralized hemispheric dysfunction in this psychotic disorder^80–82^. In line with our findings, a recent fMRI study observed a dissociable network signature of SCZ, characterized by the coexistence of preserved intra-hemispheric connectivity organization and inter-hemispheric connectivity disruptions^83^, underscoring the potential functional deficits in inter-hemispheric information exchanges in SCZ. The corpus callosum is a major white matter tract that facilitates efficient inter-hemispheric neurosignal transmission. Similar to our results, a previous MR study suggested that one standard deviation decrease in the orientation dispersion index of the forceps major, and one standard deviation increase in the mean diffusivity of the tapetum, were associated with 32% and 35% higher odds of schizophrenia risk, respectively^19^. In the reverse MR, we observed putative causal effects of SCZ on the long-range cortico-subcortical hyperconnectivity, including enhanced connectivity between the putamen and DMN, the putamen and the visual network, as well as within the amygdala-limbic circuitry. Nevertheless, the disruption landscape of the cortico-subcortical connection in SCZ reported in previous observational studies remains elusive^84–87^, partly due to inability in fully controlling for confounding factors. According to the connectome architecture of SCZ from a recent worldwide ENIGMA study^88^, our findings may be partially explained by network-spreading pathological processes propagating from subcortical epicenters (e.g., the putamen, amygdala) to distal cortical regions. Therefore, our MR results may provide novel insights to elucidate the association between structural dysconnectivity and SCZ.

Our MR analyses suggest potential causality between brain structural connectome and major psychiatric disorders. Previous MR study has examined the causal relationships between the brain functional connectome and psychiatric disorders^89^, but since functional connectivity is closely linked to and dependent on the structural connectivity^90^, investigating the causal relationships between brain structural connectome and psychiatric disorders remains of significant importance. Furthermore, two MR studies^91,92^ have explored the structural and functional connectivity within the “Yeo 7” functional networks^22^ and their causal relationships with depression. However, these studies did not examine the structural connectivity between pairs of functional networks or between functional networks and subcortical structures, nor did they address other common psychiatric disorders.

Our study has several limitations that should be acknowledged. First, the GWAS for brain structural connectome was based on cohorts from the UK Biobank. Although we meticulously selected large-scale GWAS datasets that did not include UK Biobank participants to minimize sample overlap, a potential maximum overlap rate of approximately 5.29% for AN^23^ and 2.15% for PTSD^24^ still exists. Due to the inaccessibility of detailed participants’ information, we were unable to exclude overlapping participants. Second, differences in the age distributions between the cohorts used in the GWAS for brain structural connectome and those for psychiatric disorders may introduce bias, particularly for age-related psychiatric disorders. Third, all GWAS summary data used in this study were derived from populations of European ancestry. Therefore, the generalizability of our findings to other populations requires further investigation. Fourth, while we excluded SNPs associated with common confounders during IV selection, some unmeasured confounders may still persist. Potential biases could arise from unobserved confounding factors such as population stratification and assortative mating^93^. Additionally, MR analysis relies on the principle of gene-environment equivalence, which assumes genetic variation-induced changes in exposures have identical downstream effects on outcomes as environmental changes^17,94^. However, genetic variations may not accurately mimic environmental changes. Furthermore, MR estimates reflect the lifetime effects of exposures on outcomes, which could lead to larger effect sizes compared to estimates derived from RCTs or other approaches that measure effects over specific time frames^17^. Therefore, despite the rigorous IV selection and sensitivity analyses conducted, the findings of our MR study should be interpreted with caution when considering clinical applications and warrant further validation through longitudinal clinical studies.

In conclusion, we explore the causal relationships between brain structural connectome and major psychiatric disorders by conducting bidirectional two-sample MR analyses with 206 white-matter structural connectivity phenotypes and 13 psychiatric disorders. The results shed lights on the etiology of major psychiatric disorders at the level of brain structural connectome, as well as provides insights into potential biomarkers for detection and prevention of the psychiatric disorders.

## Methods

### GWAS of brain structural connectome

We used the GWAS summary statistics of human brain structural connectome from 26,333 participants of European ancestry in the UK Biobank, processed from Wainberg et al^21^. Specifically, the density and connectivity of white-matter fibers between pairs of brain regions were quantified based on 214 predefined regions, which include 200 cortical parcels from the Schaefer atlas^95^ and 14 subcortical parcels from the Harvard-Oxford atlas. A GWAS was subsequently conducted on the brain structural connectome to investigate the associations between 206 white-matter structural connectivity measures and the 9,423,516 variants present in the imputed genotypes of the UK Biobank.

The 206 white-matter structural connectivity measures include: (1) three hemisphere-level connectivity measures, including left intra-hemisphere, right intra-hemisphere and inter-hemisphere connectivity; (2) a total of 105 cortical network-level connectivity measures, encompassing white-matter structural connectivity within the seven functional networks^22^ of both hemispheres. Specifically, 14 measures were derived from within-network connectivity, while 91 measures were derived from between-network connectivity; and (3) a total of 98 cortical-to-subcortical connectivity measures, including white-matter structural connectivity between the seven functional networks^22^ of both hemispheres and seven subcortical structures: the thalamus, caudate, putamen, pallidum, hippocampus, amygdala, and accumbens.

### GWAS of psychiatric disorders

In our study, we collected publicly available GWAS summary statistics for thirteen psychiatric disorders, including attention-deficit/hyperactivity disorder^96^ (38,691 cases and 186,843 controls), anorexia nervosa^23^ (16,992 cases and 55,525 controls), anxiety disorders^97^ (50,486 cases and 330,460 controls), autism spectrum disorder^98^ (18,381 cases and 27,969 controls), alcohol use disorder^99^ (113,325 cases and 639,923 controls), bipolar disorder^100^ (40,463 cases and 313,436 controls), cannabis use disorder^101^ (42,281 cases and 843,744 controls), depression^97^ (53,313 cases and 394,756 controls), obsessive-compulsive disorder^97^ (2,490 cases and 403,817 controls), opioid use disorder^102^ (15,251 cases and 538,935 controls), post-traumatic stress disorder^24^ (137,136 cases and 1,085,746 controls), schizophrenia^103^ (53,386 cases and 77,258 controls), and Tourette syndrome^104^ (4,819 cases and 9,488 controls). Given that the GWAS of brain structural connectivity was conducted in the European-ancestry population of the UK Biobank, we tried to avoid participants overlap between the psychiatric disorder GWAS and brain structural connectome GWAS. There was no sample overlap with the UK Biobank in 11 of the psychiatric disorders, except for anorexia nervosa and post-traumatic stress disorder. In the case of anorexia nervosa, the largest obtainable GWAS sample was utilized, exhibiting a maximum overlap rate of 5.29% with the brain structural connectome sample. Post-traumatic stress disorder was also examined using the largest accessible GWAS sample, featuring an overlap rate of approximately 2.15% and a sample size in the millions. Given these parameters, the likelihood of introducing meaningful bias into the results is considered minimal^19,25,26^. To minimize confounding due to genetic ancestry differences and other contextual factors that could inflate MR results, we restricted all psychiatric disorder GWAS datasets utilized the study to European-ancestry samples. Detailed information on these GWAS samples is summarized in Table 1 and Supplementary Table 2.

### Selection of IVs

For MR analysis to be valid, three fundamental assumptions must be satisfied: (1) the IVs must be strongly associated with the exposure; (2) the IVs must be independent of confounders that influence both the exposure and the outcome; and (3) the IVs must affect the outcome only through the exposure, without exerting any direct effect on the outcome or through alternative pathways, which is also referred to as the absence of horizontal pleiotropy^27^.

To satisfy these assumptions, we first selected SNPs that are strongly associated with the exposure (pval < 5e-8). Clumping was then performed using the “TwoSampleMR” R package^105^, with parameters r² = 0.001 and a window size of 1,000 kb, using the 1000 Genomes European data as the reference panel. Incompatible SNPs (those that do not follow the principle of complementary base pairing) were checked and removed, and palindromic SNPs with a minor allele frequency close to 0.5 were excluded to avoid potential ambiguity. The harmonization procedure was carried out using the “TwoSampleMR” R package^105^ to ensure that the SNPs in the exposure and outcome GWAS summary statistics are consistent and came from the same DNA strand. Afterwards, SNPs associated with the outcome were removed (pval < 5e-5), and the Steiger test^106^ was performed to reduce the risk of potential reverse causality. Previous studies have shown that factors such as income^107,108^, education^109,110^, smoking^111,112^ and drinking^113,114^ may influence both brain structural connectome and psychiatric disorders. Therefore, we used the NHGRI-EBI Catalog^115^ (https://www.ebi.ac.uk/gwas/home) to search for and remove SNPs associated with potential confounders (pval < 5e-5). Quality control was conducted to enhance the robustness of the IVs. We used the ‘IVW_radial’ method from the “RadialMR” R package^116^ to perform Cochran’s Q test for the IVW model, and outliers were removed (pval < 5e-5).

The F statistic was calculated to assess the strength of the IVs using the following formula^117,118^,

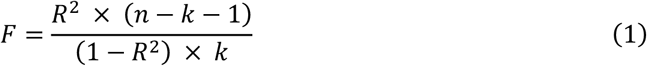

Where *R*^2^ (the variance in the exposure explained by the IVs), *n* (the sample size of the exposure GWAS), and *k* (the number of IVs) are parameters in the calculation. The *R*^2^ statistic can be calculated using the following formula^117,118^,

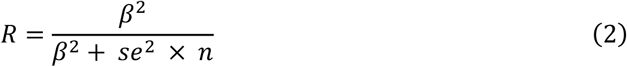

where β (effect size of the exposure) and *se* (standard error of the exposure) are parameters in the calculation. For multiple IVs, the *R*^2^ is the sum of *R*^2^ of each individual IV.

### Bidirectional MR analyses

Bidirectional two-sample MR analyses were conducted to investigate the causal relationships between brain structural connectome and psychiatric disorders. In the forward MR analyses, the brain structural connectome served as the exposure, with psychiatric disorders as the outcome. Conversely, in the reverse MR analyses, psychiatric disorders were considered the exposure, and the brain structural connectome was regarded as the outcome.

The inverse-variance weighted (IVW) regression with multiplicative random effects was applied as the primary causal inference method due to its highest statistical efficiency^119^. However, the IVW method may yield biased results if there is an average pleiotropic effect that deviates from zero. Therefore, six additional MR methods were employed to strengthen the robustness of our findings. The MR-Egger method, particularly useful when there is directional pleiotropy among IVs, provides a consistent causal effect estimate through its slope^120^. The weighted median method yields reliable causal effects though with invalid IVs under the assumption that at least half of the IVs are valid^121^. The weighted mode method groups IVs based on similar causal effects and provides consistent estimates if the majority of IVs in the largest cluster are valid^30^. MR-robust adjusted profile scores (MR-RAPS) accounts for systematic and idiosyncratic pleiotropy and enables robust causal inference with many weak IVs^122^. The contamination mixture method offers reliable estimates for causal analysis using hundreds of IVs with the presence of invalid IVs^123^. Debiased inverse variance weighted (Debiased IVW) effectively reduces bias related to weak IVs and enhances robustness in situations with multiple weak IVs^124^. All the above methods were conducted using the “TwoSampleMR”^105^, “mr.raps”^122^ and “MendelianRandomization”^125^ R package.

To account for multiple comparisons, our study applied a Bonferroni-corrected significance threshold of 1.2316 × 10^−4^ (0.05/206/2, 206 denotes the number of white-matter structural connectivity phenotypes, 2 represents bidirectional MR analyses). Additionally, we set a nominal significance threshold of 1 × 10^−3^, considering that results slightly below the Bonferroni threshold may still be of suggestive value. To make sure our MR analyses solid, we only expect causal links that meet the following criteria: there are sufficient SNPs for sensitivity analyses (i.e., at least four IVs)^89^, the directions of estimates from different MR methods are consistent, and the p-value from the IVW method is below the nominal significance threshold. In addition, we calculate statistical power for the discovered causal associations using an online web tool (https://sb452.shinyapps.io/power/)^126^.

Considering that the white-matter structural connectivity phenotypes of the brain structural connectome are continuous, while psychiatric disorder phenotypes are categorical, we utilized odds ratios (OR) and beta to quantify the effect sizes in the forward and reverse MR analyses, respectively. Furthermore, we adhered to the Strengthening the Reporting of Mendelian Randomization Studies (STROBE-MR) guidelines^127^ (see Supplementary Note for details).

### Sensitivity analyses

A series of sensitivity analyses were conducted to further verify the significant MR results. First, we employed MR-Egger intercept test^120^ to detect potential directional pleiotropy (P < 0.05). Then, we performed MR-PRESSO global test^128^ to detect potential bias of horizontal pleiotropy (P < 0.05). Additionally, Cochran’s Q test^129^ was utilized to assess heterogeneity in the causal estimates across different IVs (P < 0.05). Finally, we conducted leave-one-out analyses to identify whether any single IV is disproportionately influencing the results or whether the causal effects remain consistent when each IV is excluded^17^. The MR-PRESSO global test was conducted with the “MR-PRESSO” R package^128^, while all other analyses were performed using the “TwoSampleMR” R package^105^.

## Data Availability

All GWAS summary statistics utilized in this study are publicly available. The GWAS summary statistics of 206 brain white-matter structural connectivity (GCST90302648 ∼ GCST90302853) can be downloaded from GWAS Catalog (https://www.ebi.ac.uk/gwas/). The GWAS summary statistics for ANX, Depression, and OCD are obtained from the FINNGEN R11 release (To access this data, please follow the FINNGEN R11 release guidance: https://finngen.gitbook.io/documentation/data-download). The GWAS summary statistics for the three substance use disorders (AUD, CUD, OUD) can be accessed at https://medicine.yale.edu/lab/gelernter/stats/. Summary statistics for other psychiatric disorders are available from the PGC (https://pgc.unc.edu/for-researchers/download-results/). The GWAS summary statistics for birth length, used as a negative control, can be downloaded from https://www.decode.com/summarydata/.

## Acknowledgements

This work was supported by National Natural Science Foundation of China (grant No. 62376068, 62350710797, 32361143787), by Guangdong Basic and Applied Basic Research Foundation (grant 2023A1515010792, No. 2023B1515120065), by Guangdong S&T programme (grant No. 2023A0505050109), by Shenzhen Science and Technology Innovation Program (grant No. JCYJ20220818102414031, GXWD20231129121139001, JCYJ20240813110522029). We would like to thank all the participants and investigators for contributing to the related GWAS summary data.

## Ethics committee approval and participant informed consent

Since all the GWAS summary data utilized in this study are publicly available, no additional ethical approval or participant consent was necessary. Details regarding ethical approval and participant consent can be found in the respective original GWAS publications.

