## Supplementary Figures for "Mendelian randomization analyses uncover causal relationships between brain structural connectome and risk of psychiatric disorders"

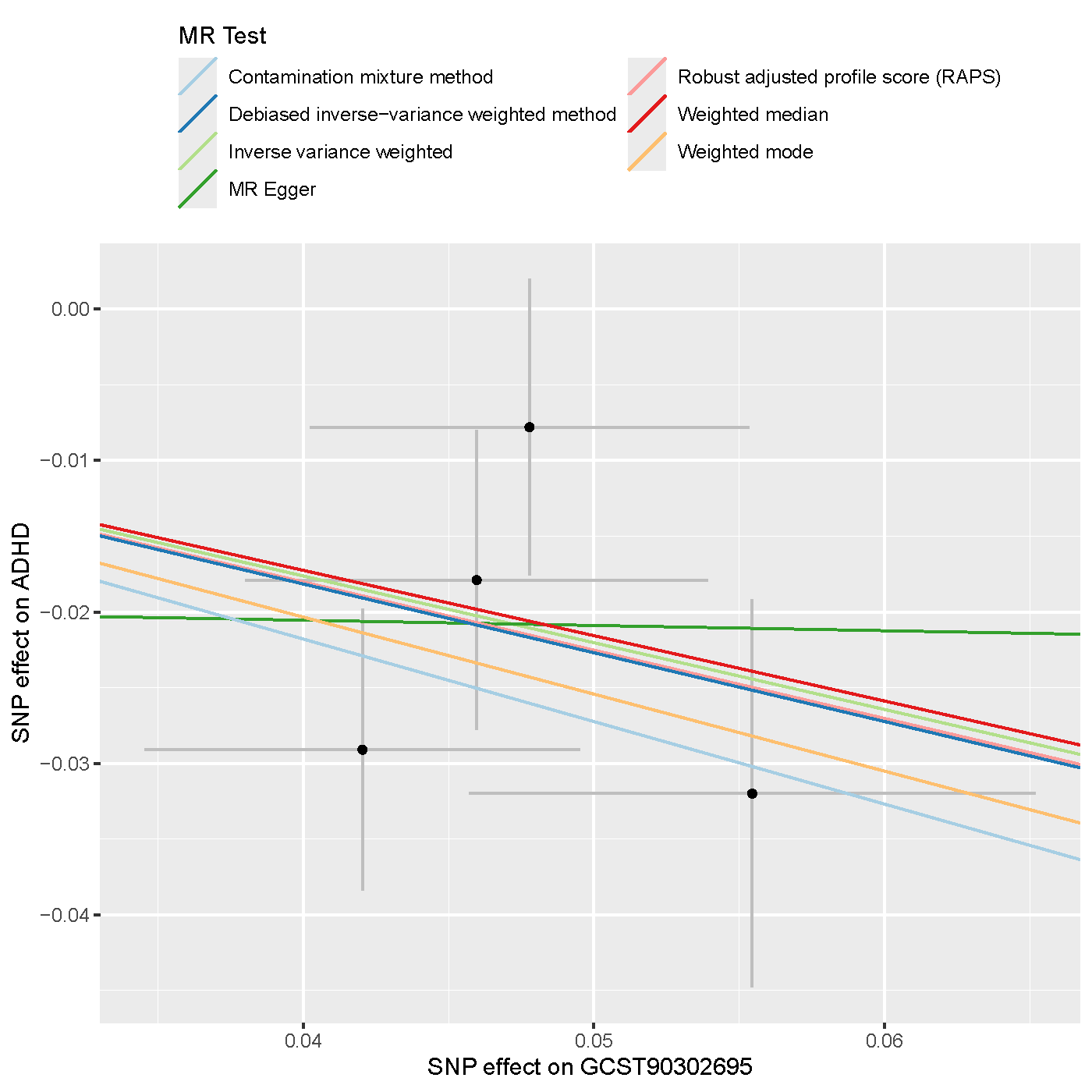

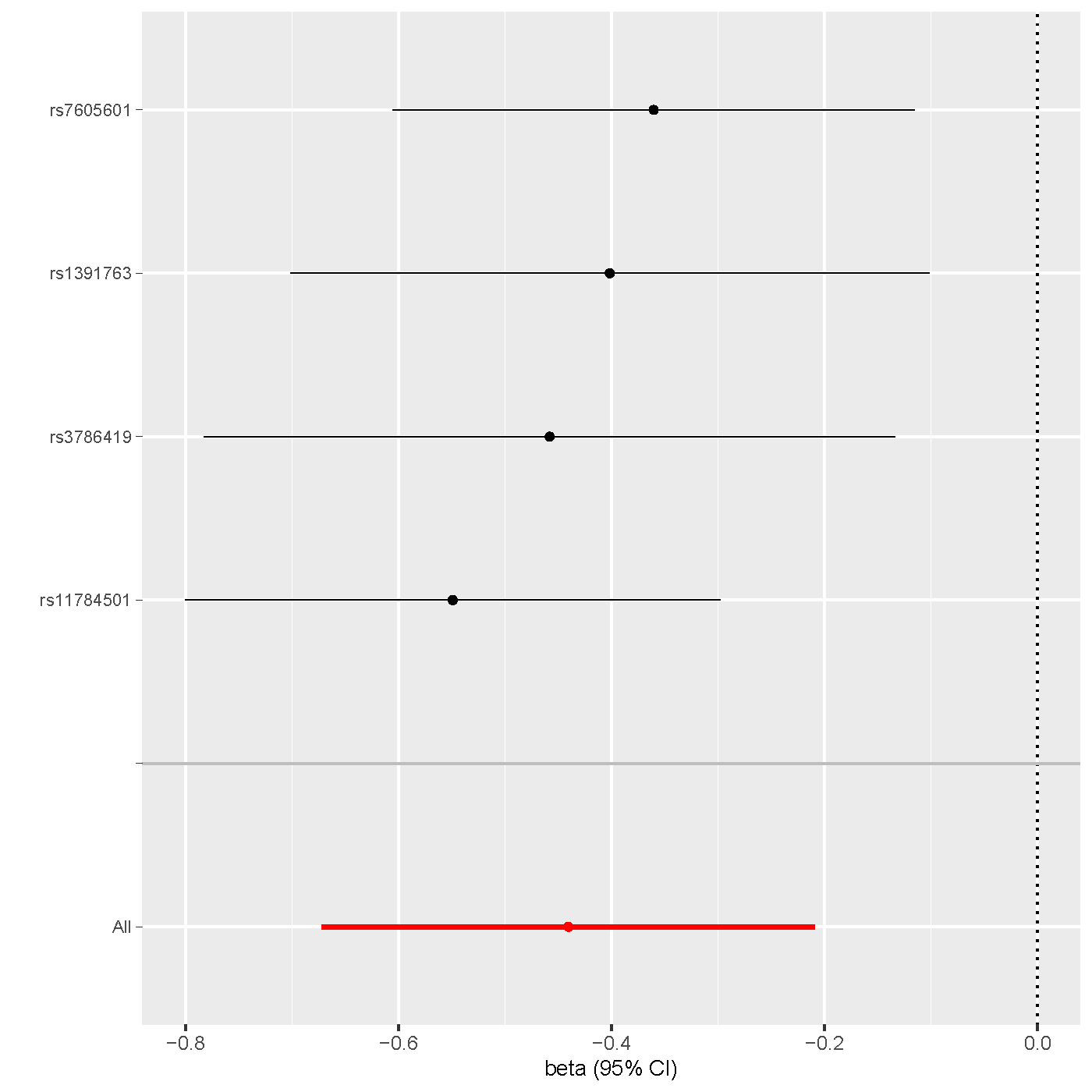

**Supplementary Figure 1 | Left-hemisphere dorsal attention network to right-hemisphere somatomotor network white-matter structural connectivity (GCST90302695) & ADHD**. **Left**: scatter plot displaying genetic associations with GCST90302695 (x-axis) over genetic associations with ADHD (y-axis) across seven different MR methods. **Right**: leave-one-out analysis plot of IVW results for GCST90302695-ADHD exposure-outcome pair identified in forward MR analyses, showing effect estimates for all SNPs and after excluding each SNP individually (y-axis), with beta (log odds ratio) and 95% confidence intervals (CI) on the x-axis.

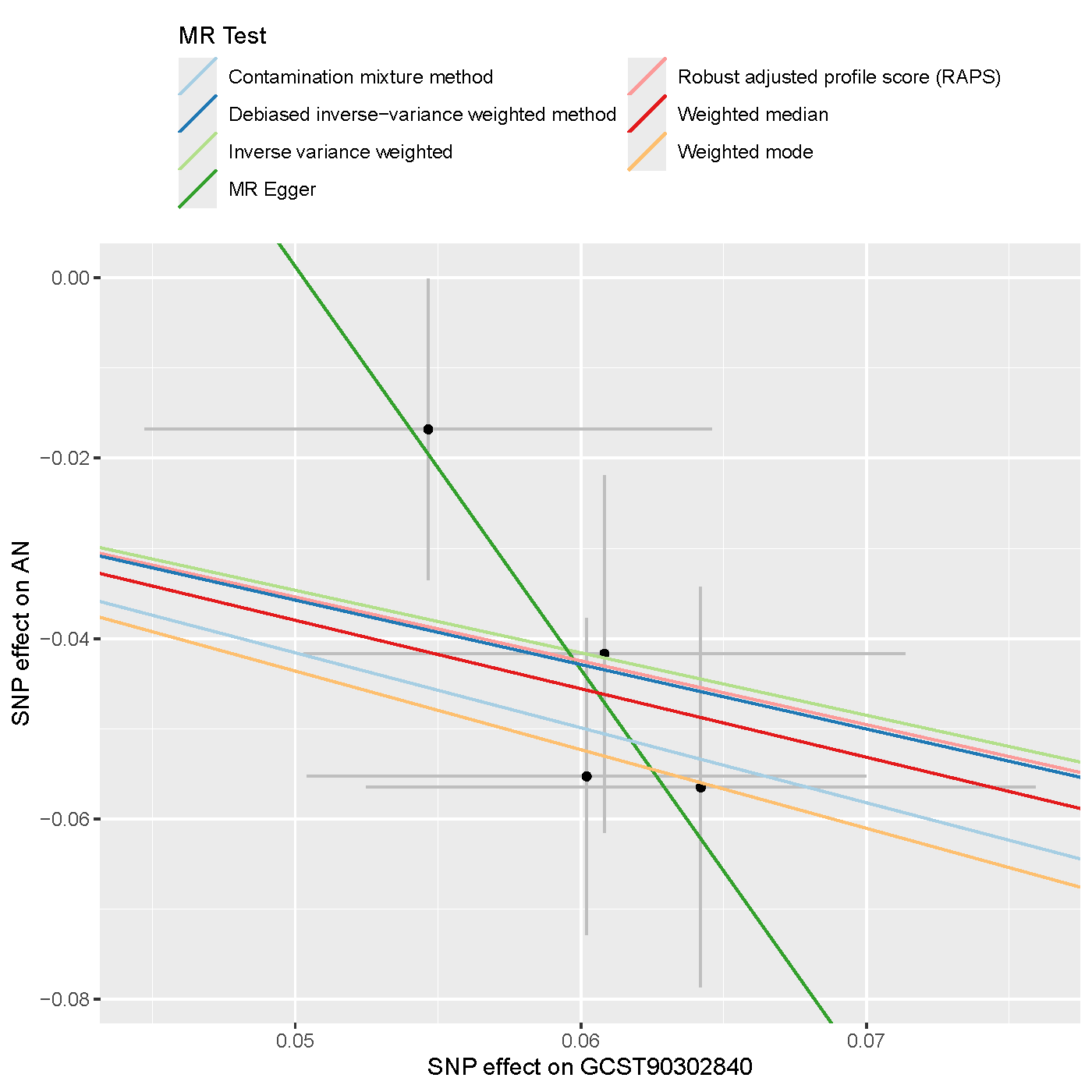

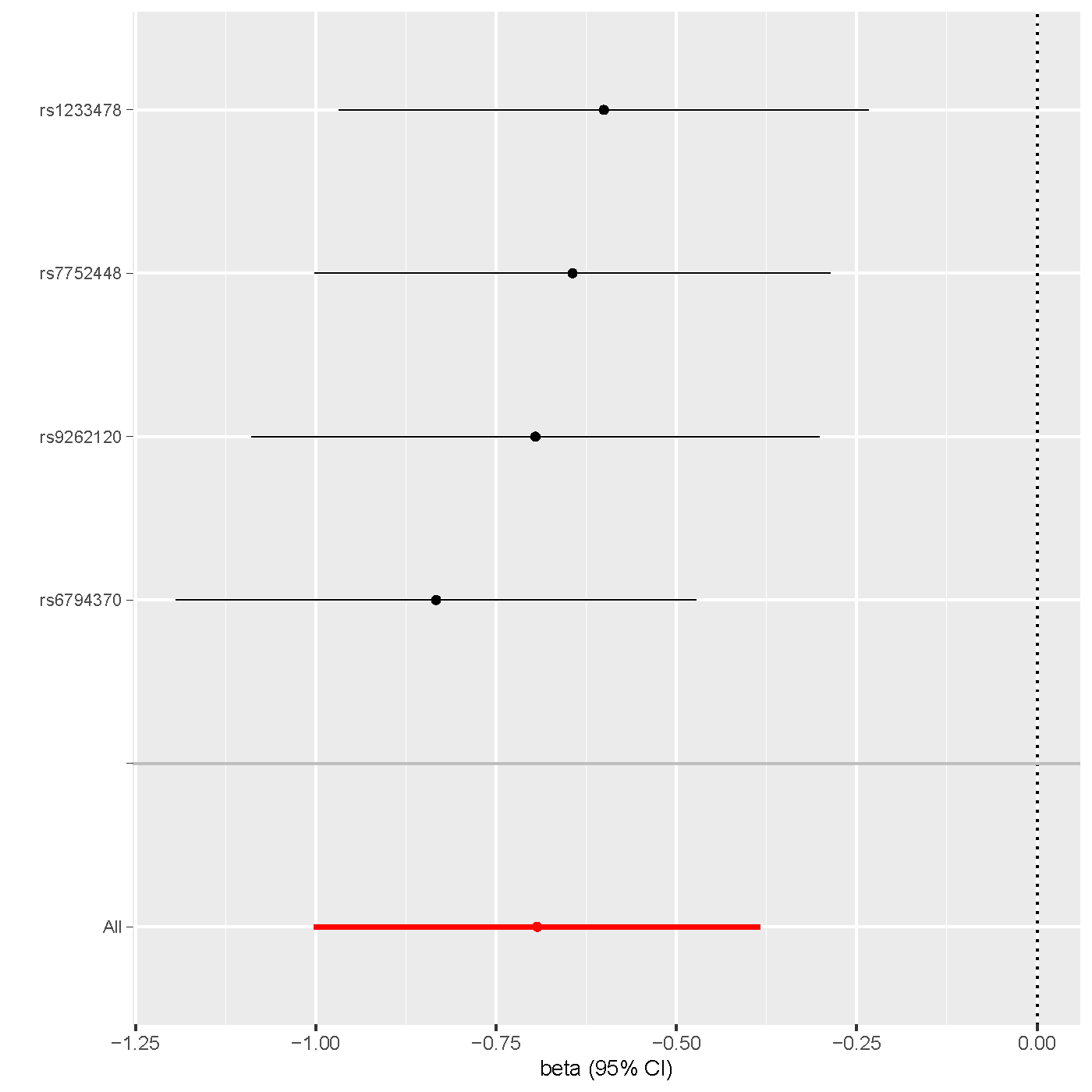

**Supplementary Figure 2 | Right-hemisphere control network to hippocampus white-matter structural connectivity (GCST90302840) & AN**. **Left**: scatter plot displaying genetic associations with GCST90302840 (x-axis) over genetic associations with AN (y-axis) across seven different MR methods. **Right**: leave-one-out analysis plot of IVW results for GCST90302840-AN exposure-outcome pair identified in forward MR analyses, showing effect estimates for all SNPs and after excluding each SNP individually (y-axis), with beta (log odds ratio) and 95% confidence intervals (CI) on the x-axis.

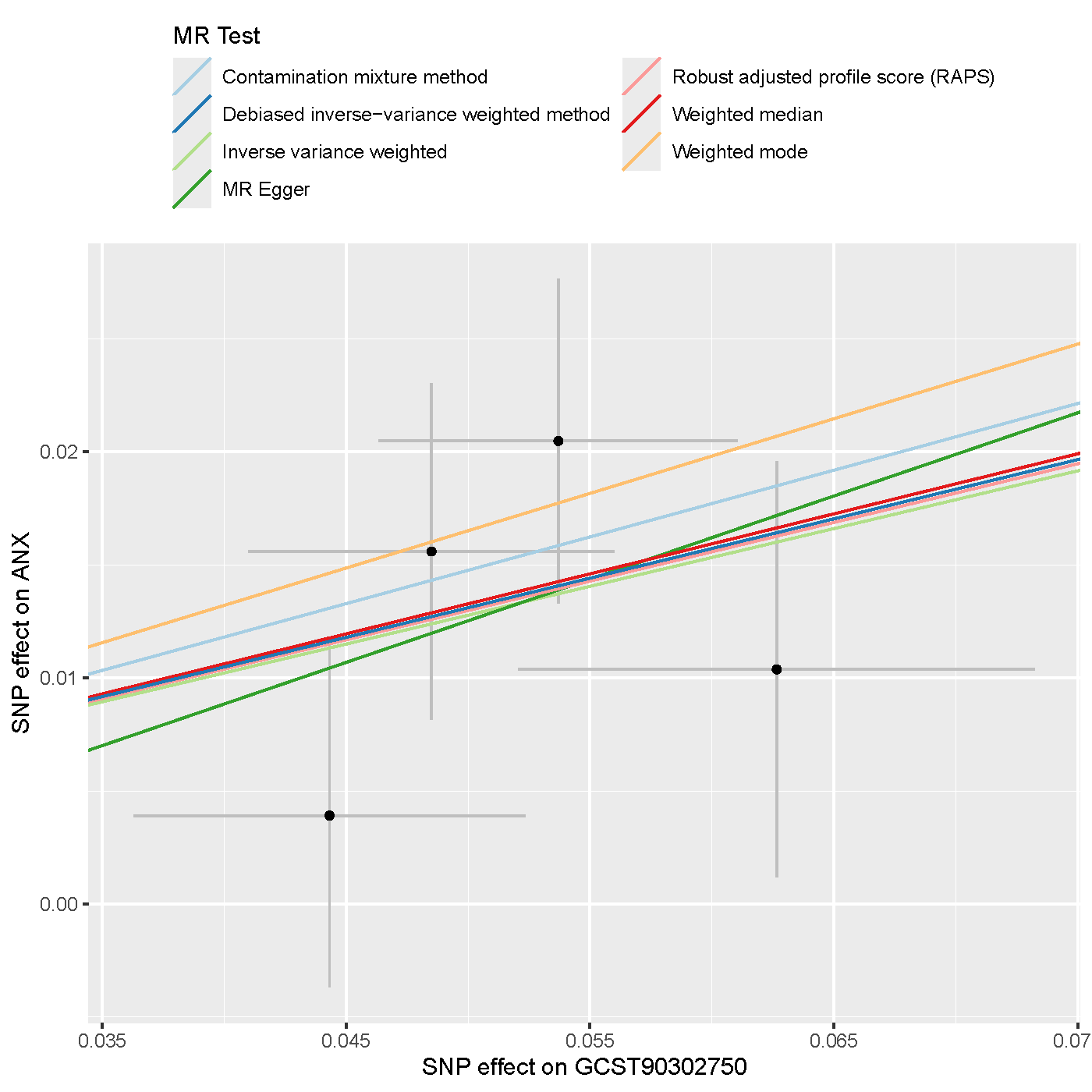

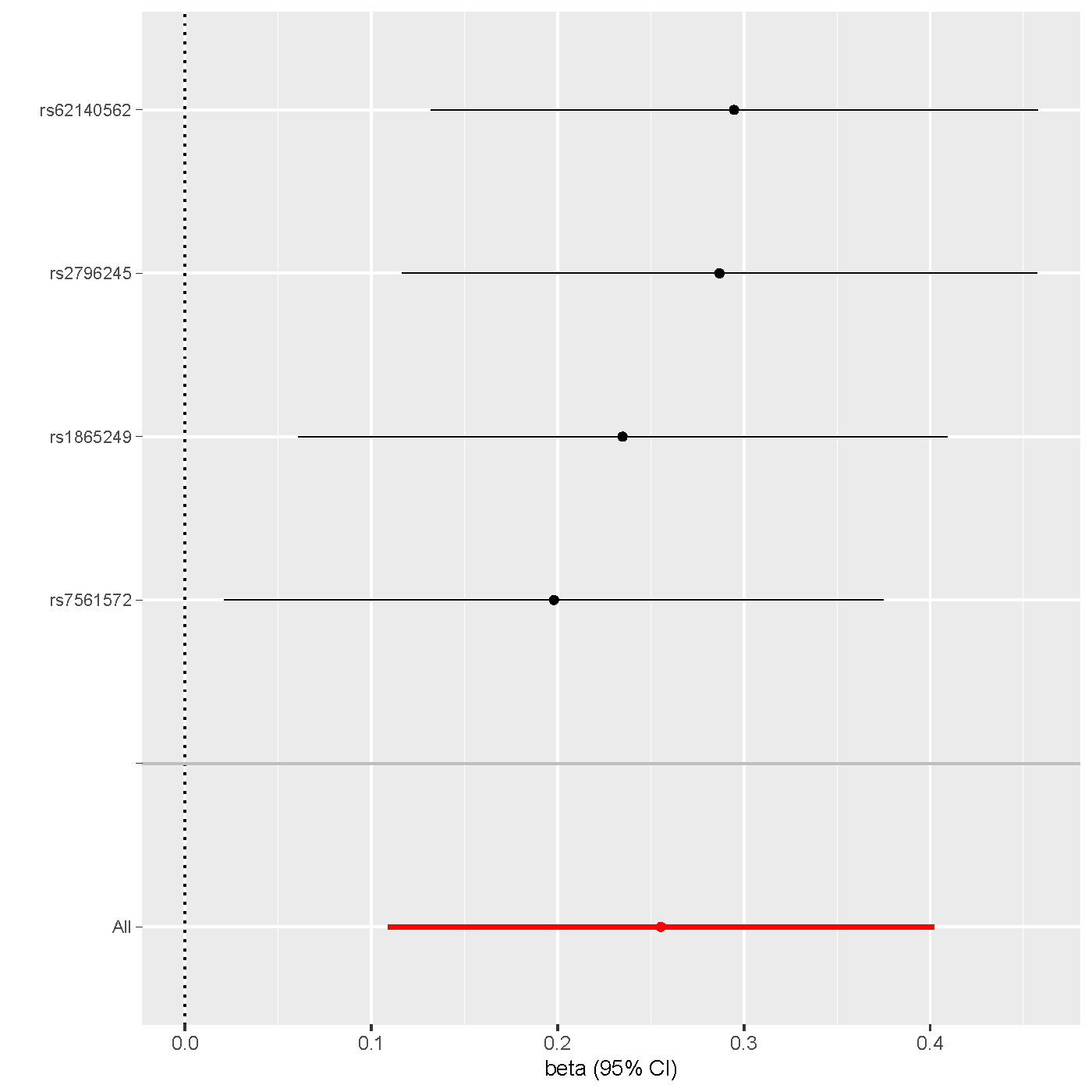

**Supplementary Figure 3 | Left-hemisphere control network to right-hemisphere control network white-matter structural connectivity (GCST90302750) & ANX**. **Left**: scatter plot displaying genetic associations with GCST90302750 (x-axis) over genetic associations with ANX (y-axis) across seven different MR methods. **Right**: leave-one-out analysis plot of IVW results for GCST90302750-ANX exposure-outcome pair identified in forward MR analyses, showing effect estimates for all SNPs and after excluding each SNP individually (y-axis), with beta (log odds ratio) and 95% confidence intervals (CI) on the x-axis.

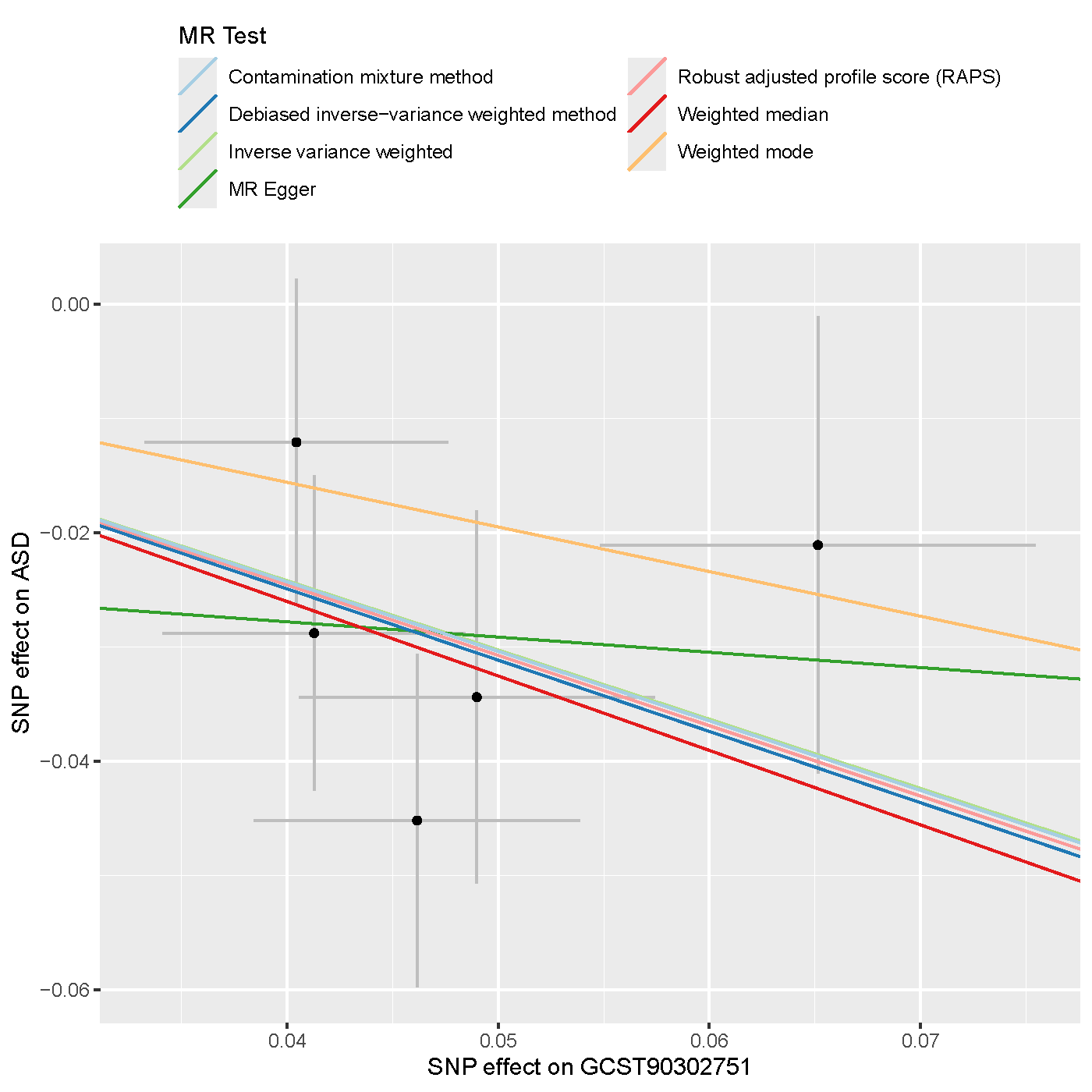

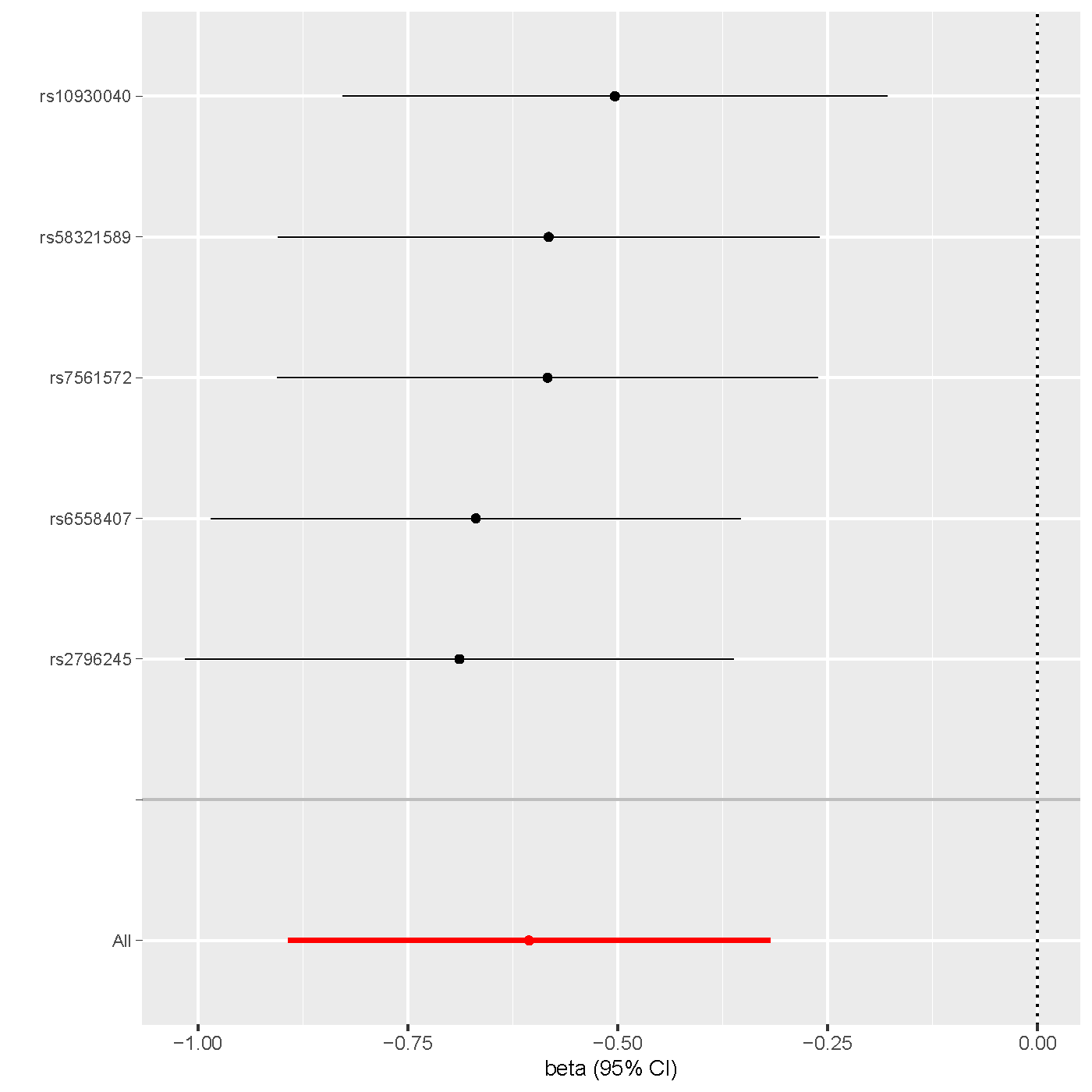

**Supplementary Figure 4 | Left-hemisphere control network to right-hemisphere default mode network white-matter structural connectivity (GCST90302751) & ASD**. **Left**: scatter plot displaying genetic associations with GCST90302751 (x-axis) over genetic associations with ASD (y-axis) across seven different MR methods. **Right**: leave-one-out analysis plot of IVW results for GCST90302751-ASD exposure-outcome pair identified in forward MR analyses, showing effect estimates for all SNPs and after excluding each SNP individually (y-axis), with beta (log odds ratio) and 95% confidence intervals (CI) on the x-axis.

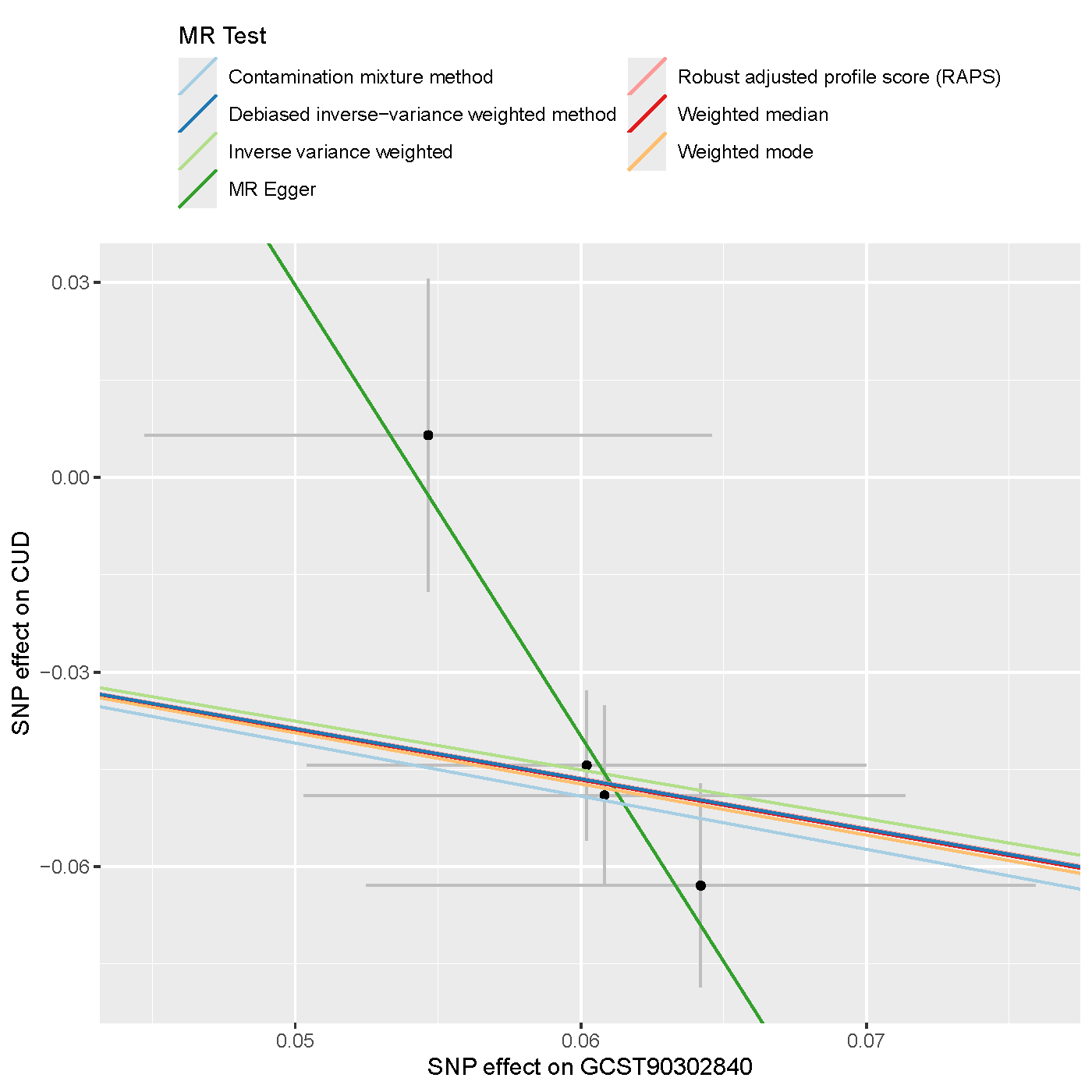

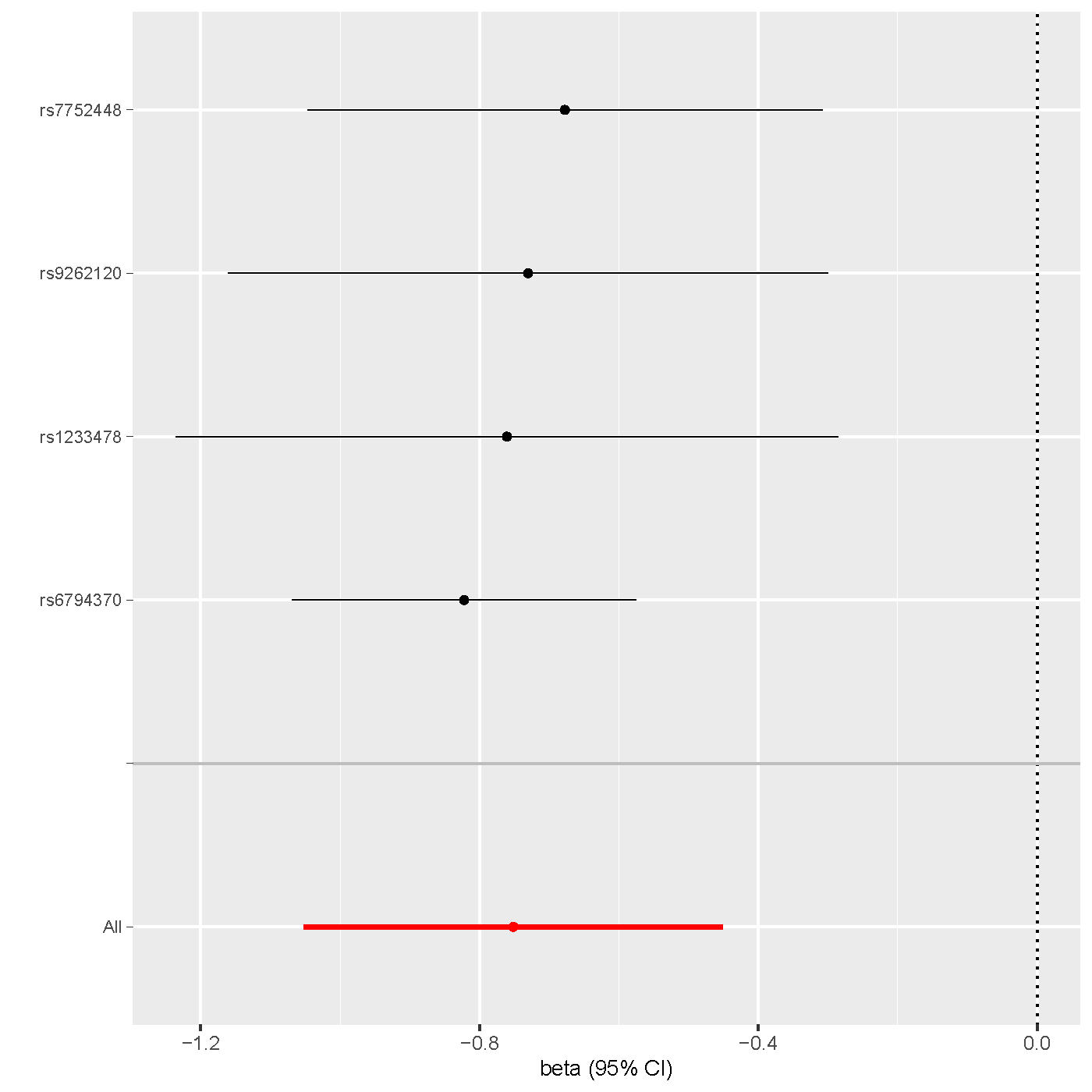

**Supplementary Figure 5 | Right-hemisphere control network to hippocampus white-matter structural connectivity (GCST90302840) & CUD**. **Left**: scatter plot displaying genetic associations with GCST90302840 (x-axis) over genetic associations with CUD (y-axis) across seven different MR methods. **Right**: leave-one-out analysis plot of IVW results for GCST90302840-CUD exposure-outcome pair identified in forward MR analyses, showing effect estimates for all SNPs and after excluding each SNP individually (y-axis), with beta (log odds ratio) and 95% confidence intervals (CI) on the x-axis.

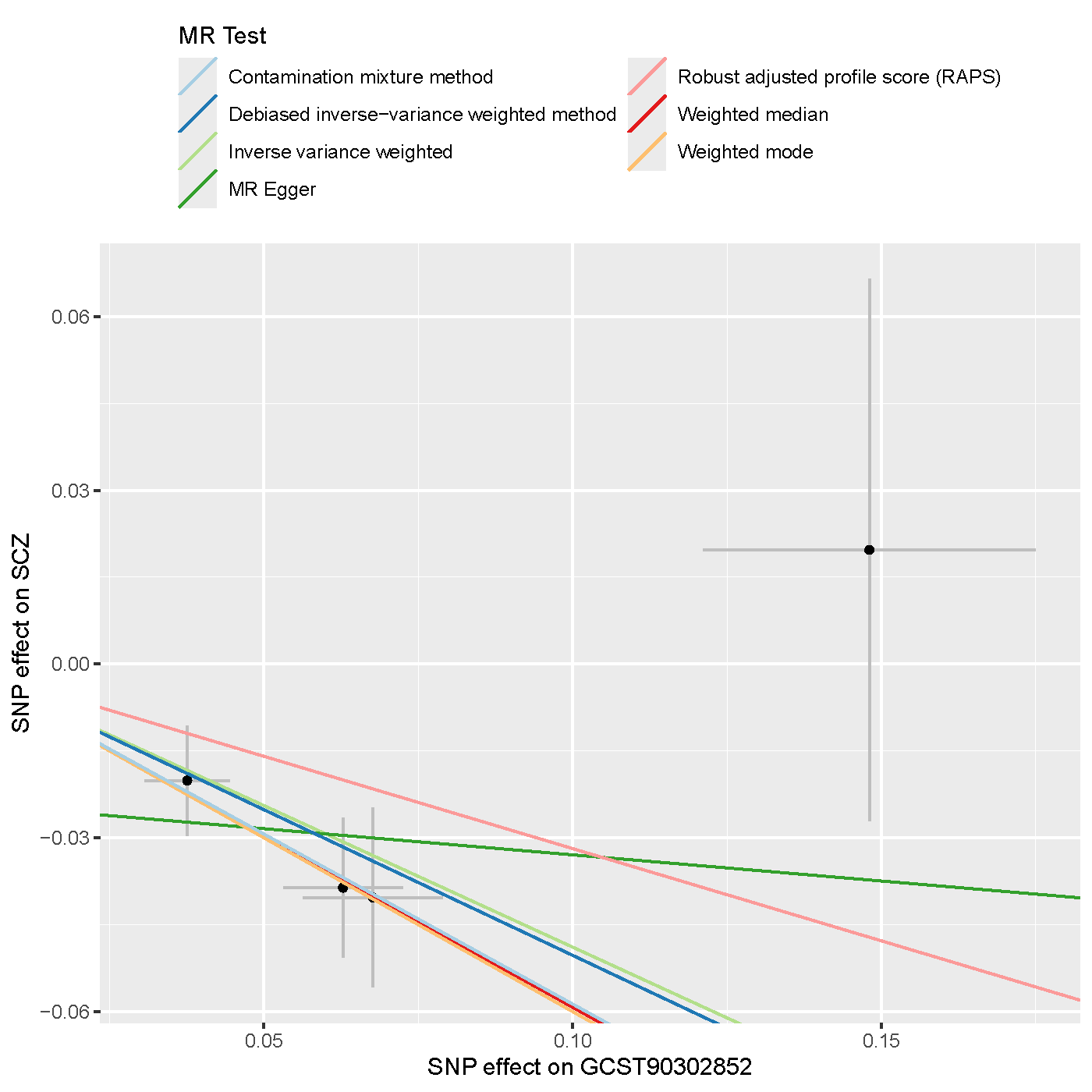

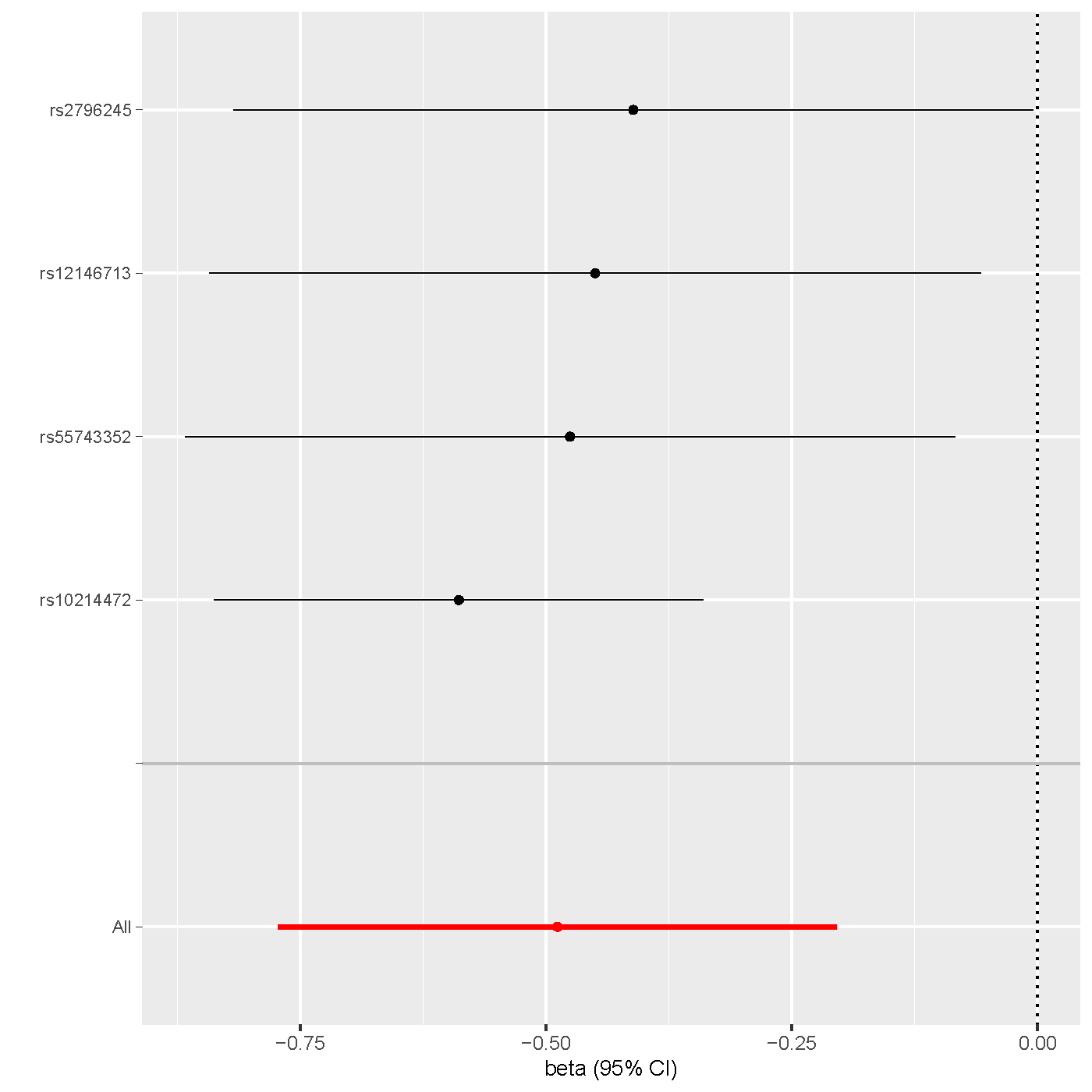

**Supplementary Figure 6 | Cross-hemisphere white-matter structural connectivity (GCST90302852) & SCZ**. **Left**: scatter plot displaying genetic associations with GCST90302852 (x-axis) over genetic associations with SCZ (y-axis) across seven different MR methods. **Right**: leave-one-out analysis plot of IVW results for GCST90302852-SCZ exposure-outcome pair identified in forward MR analyses, showing effect estimates for all SNPs and after excluding each SNP individually (y-axis), with beta (log odds ratio) and 95% confidence intervals (CI) on the x-axis.

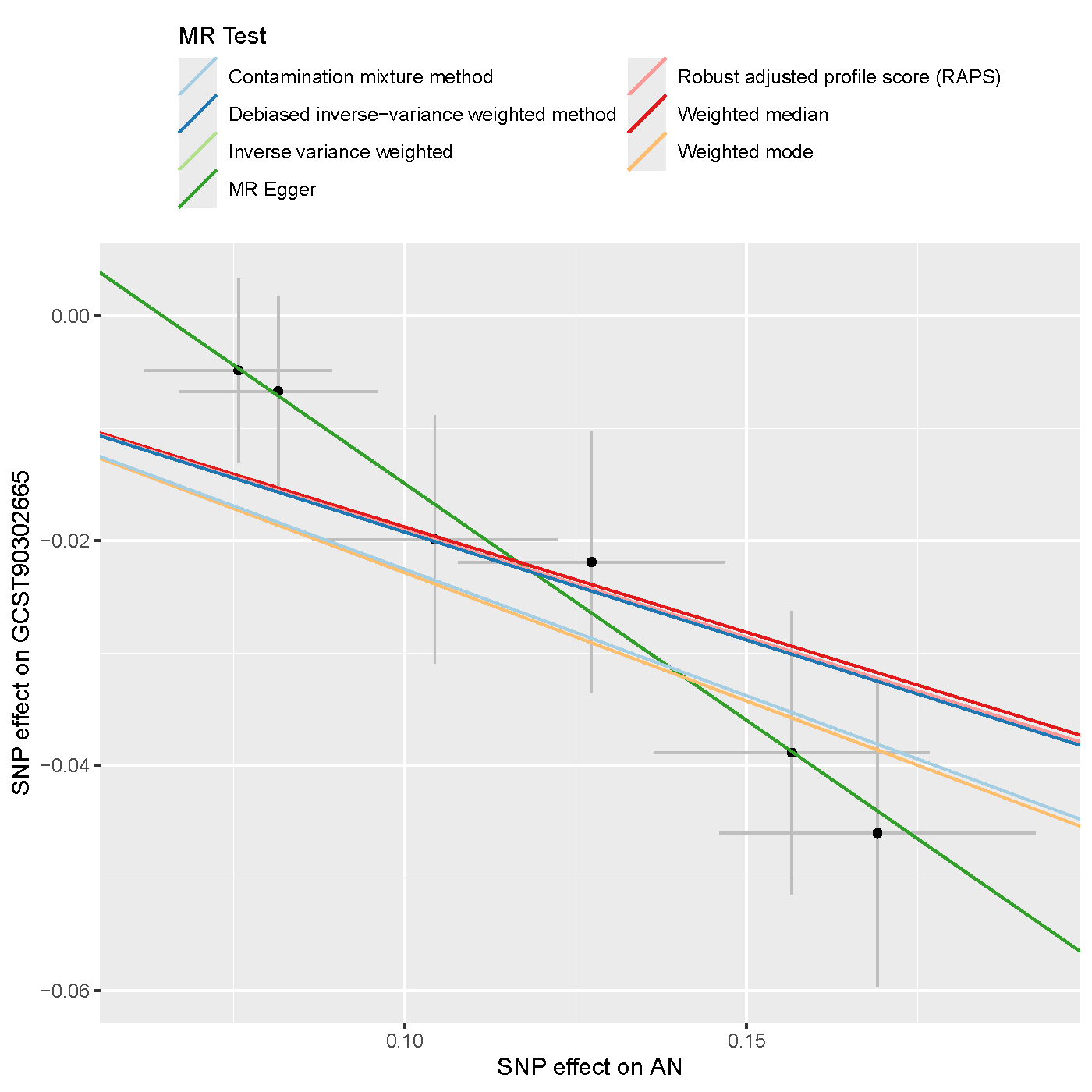

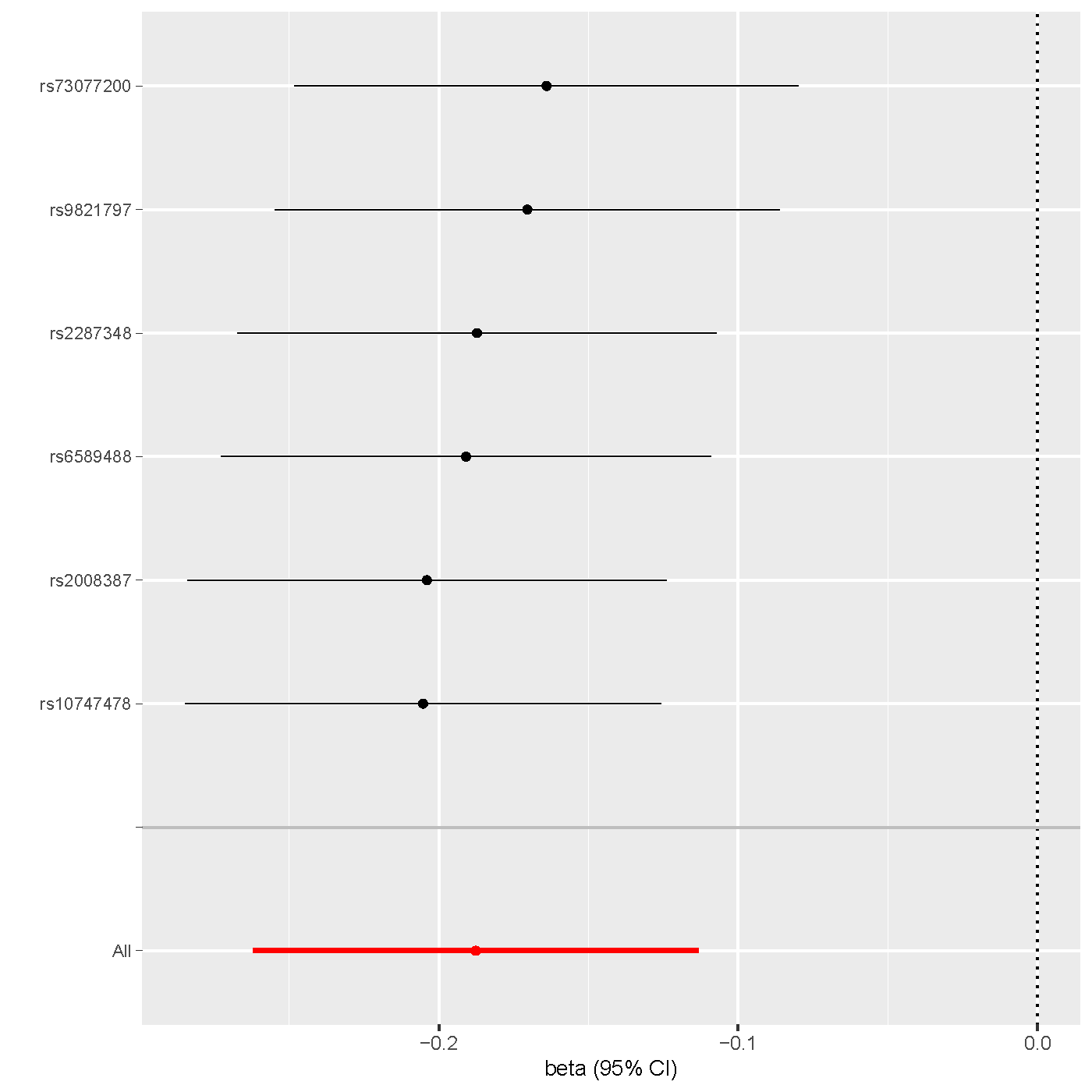

**Supplementary Figure 7 | AN & Left-hemisphere visual network to pallidum white-matter structural connectivity (GCST90302665)**. **Left**: scatter plot displaying genetic associations with AN (x-axis) over genetic associations with GCST90302665 (y-axis) across seven different MR methods. **Right**: leave-one-out analysis plot of IVW results for AN-GCST90302665 exposure-outcome pair identified in reverse MR analyses, showing effect estimates for all SNPs and after excluding each SNP individually (y-axis), with beta (log odds ratio) and 95% confidence intervals (CI) on the x-axis.

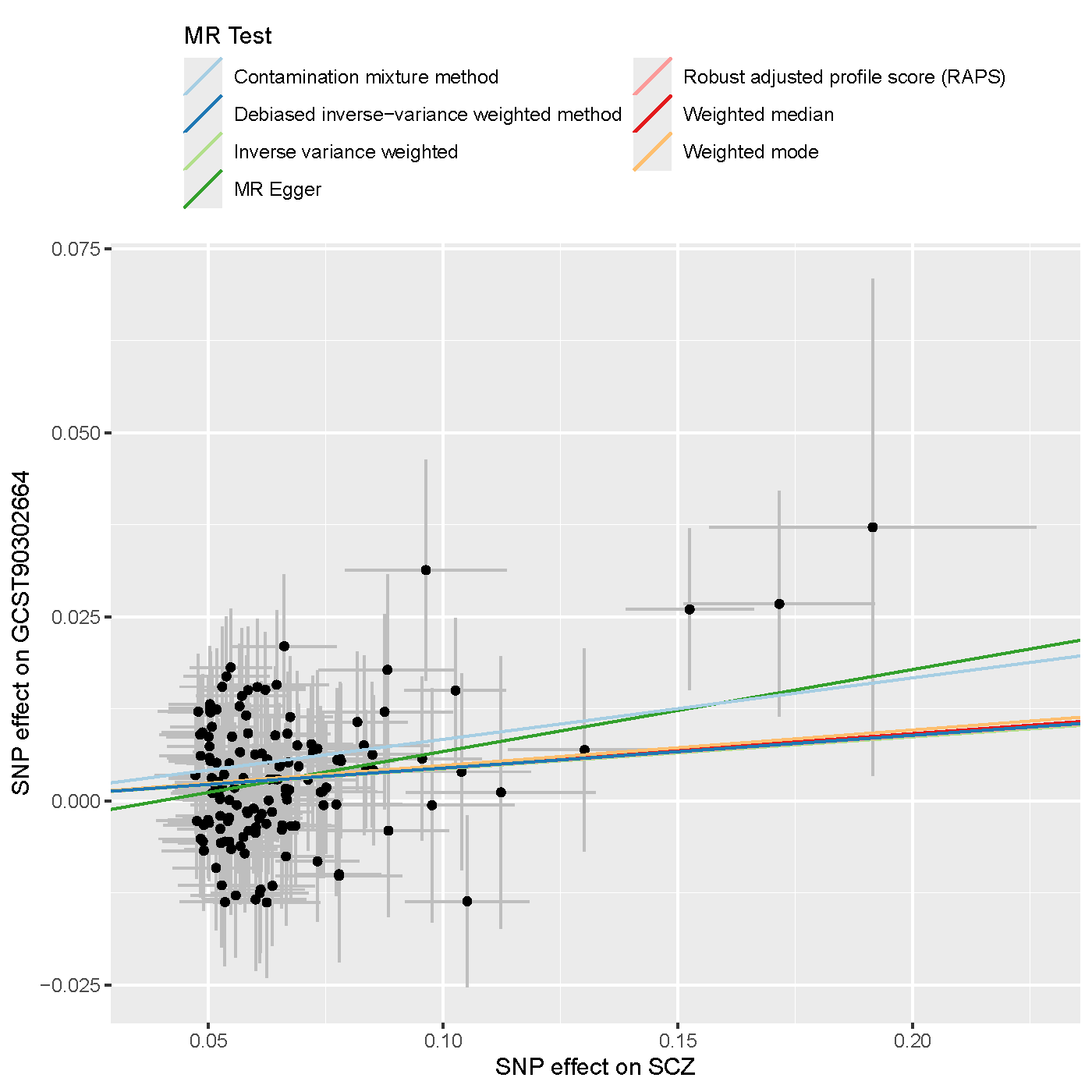

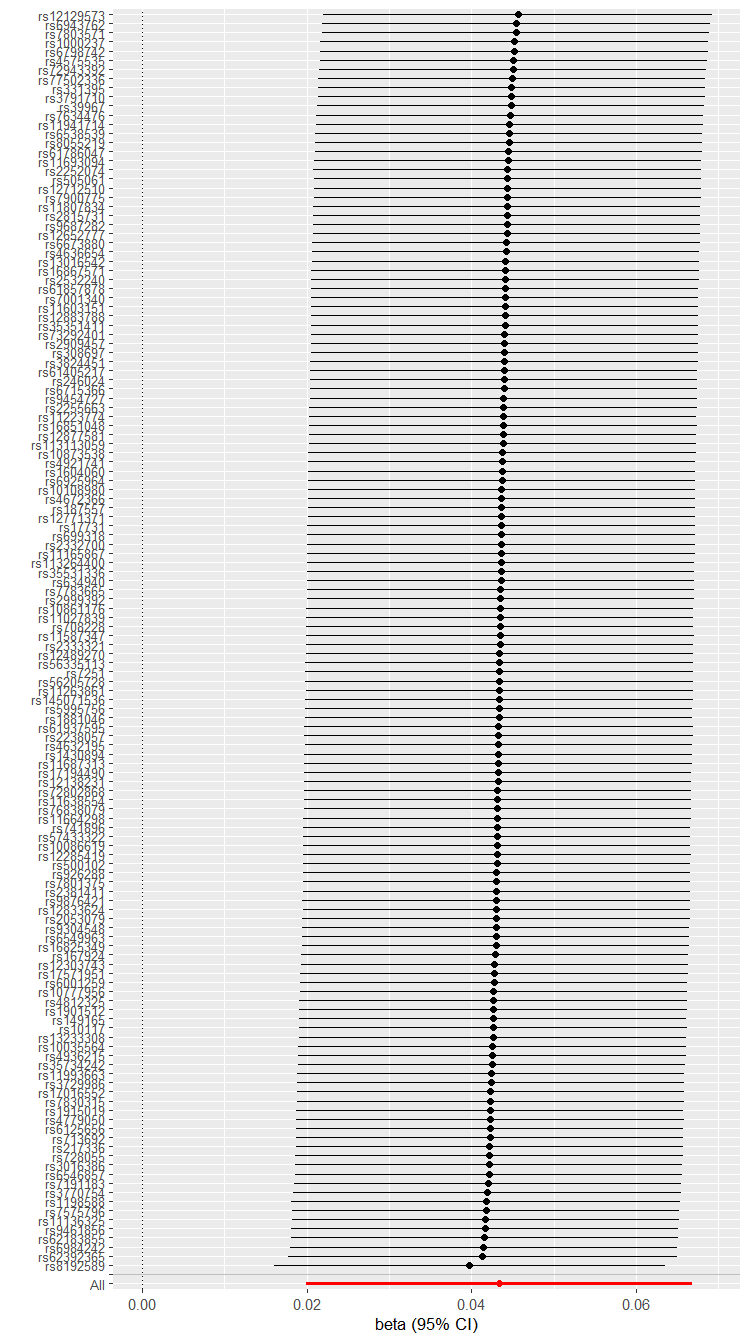

**Supplementary Figure 8 | SCZ & Left-hemisphere default mode network to putamen white-matter structural connectivity (GCST90302664)**. **Left**: scatter plot displaying genetic associations with SCZ (x-axis) over genetic associations with GCST90302664 (y-axis) across seven different MR methods. **Right**: leave-one-out analysis plot of IVW results for SCZ-GCST90302664 exposure-outcome pair identified in reverse MR analyses, showing effect estimates for all SNPs and after excluding each SNP individually (y-axis), with beta (log odds ratio) and 95% confidence intervals (CI) on the x-axis.

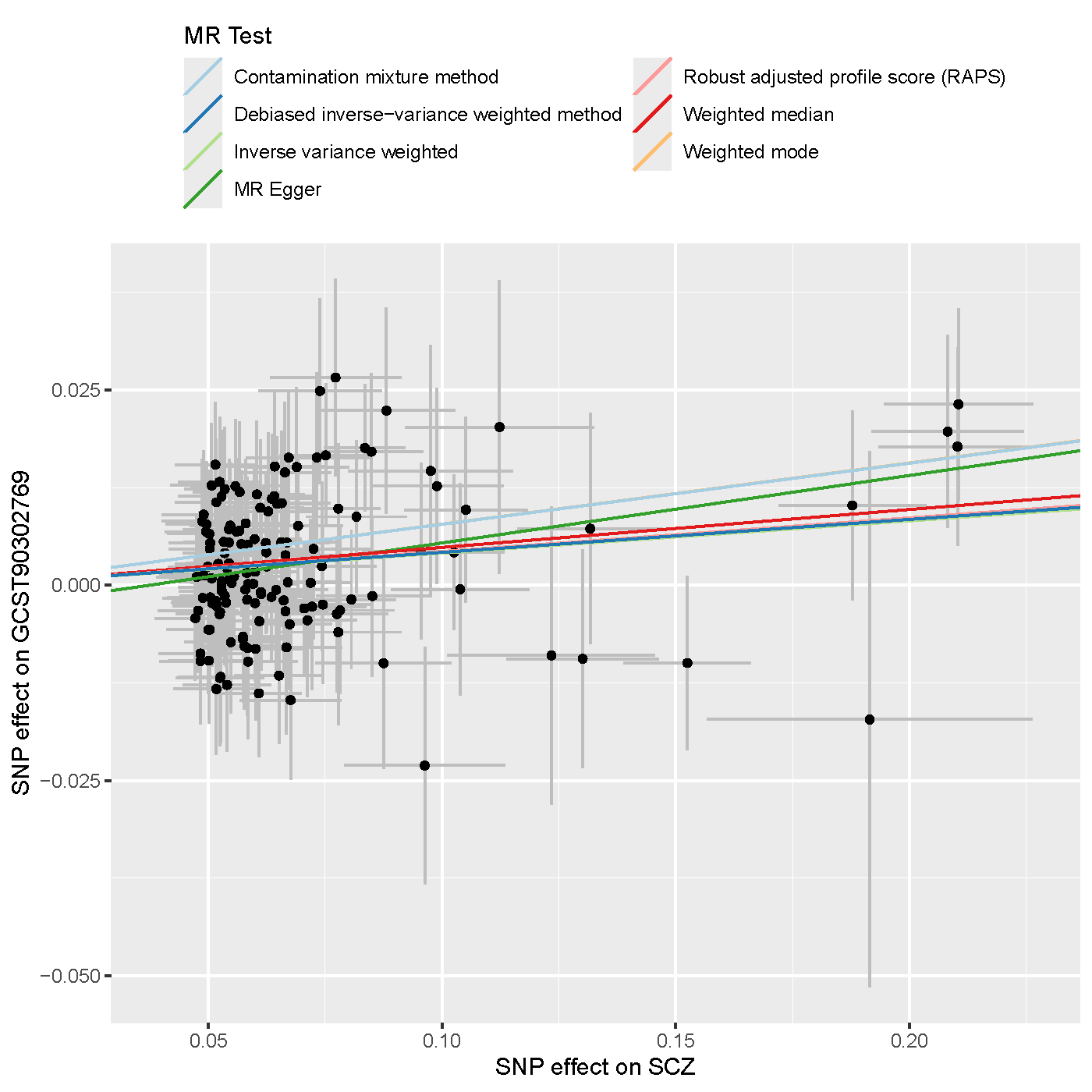

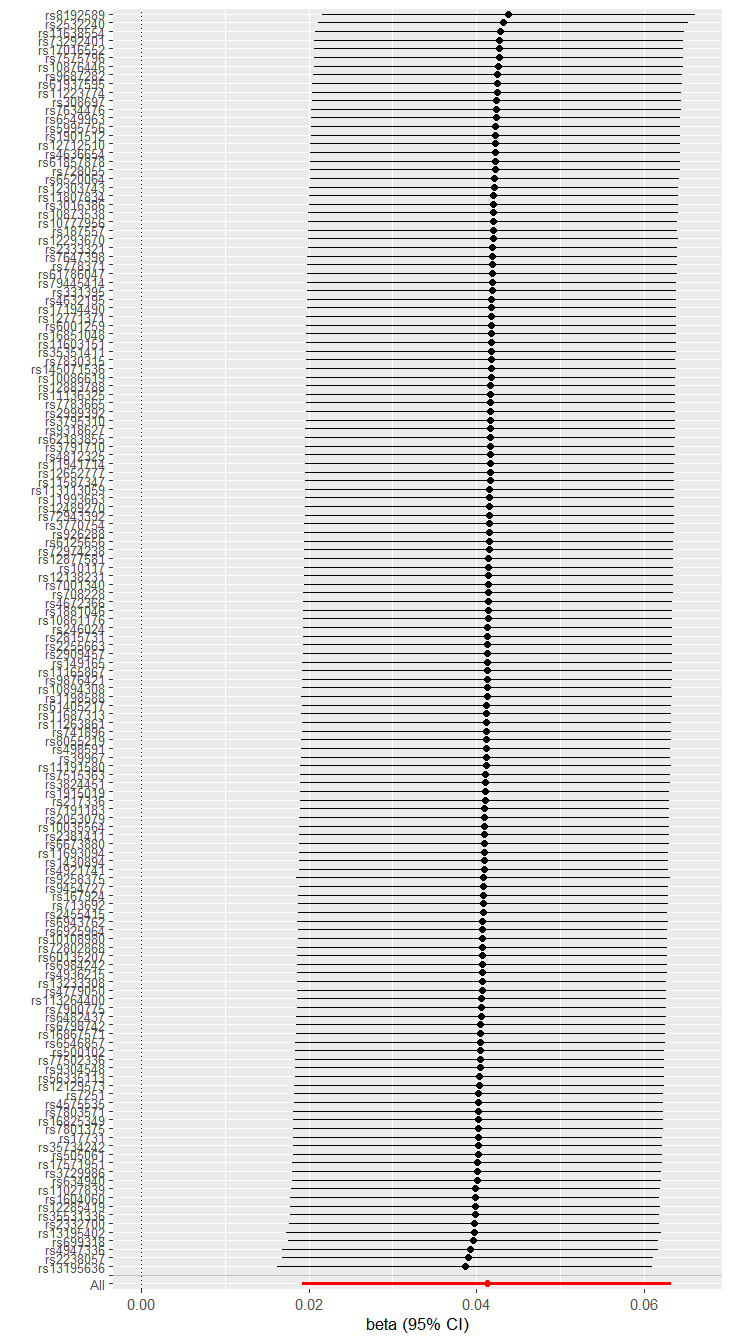

**Supplementary Figure 9 | SCZ & Left-hemisphere visual network to putamen white-matter structural connectivity (GCST90302769)**. **Left**: scatter plot displaying genetic associations with SCZ (x-axis) over genetic associations with GCST90302769 (y-axis) across seven different MR methods. **Right**: leave-one-out analysis plot of IVW results for SCZ-GCST90302769 exposure-outcome pair identified in reverse MR analyses, showing effect estimates for all SNPs and after excluding each SNP individually (y-axis), with beta (log odds ratio) and 95% confidence intervals (CI) on the x-axis.

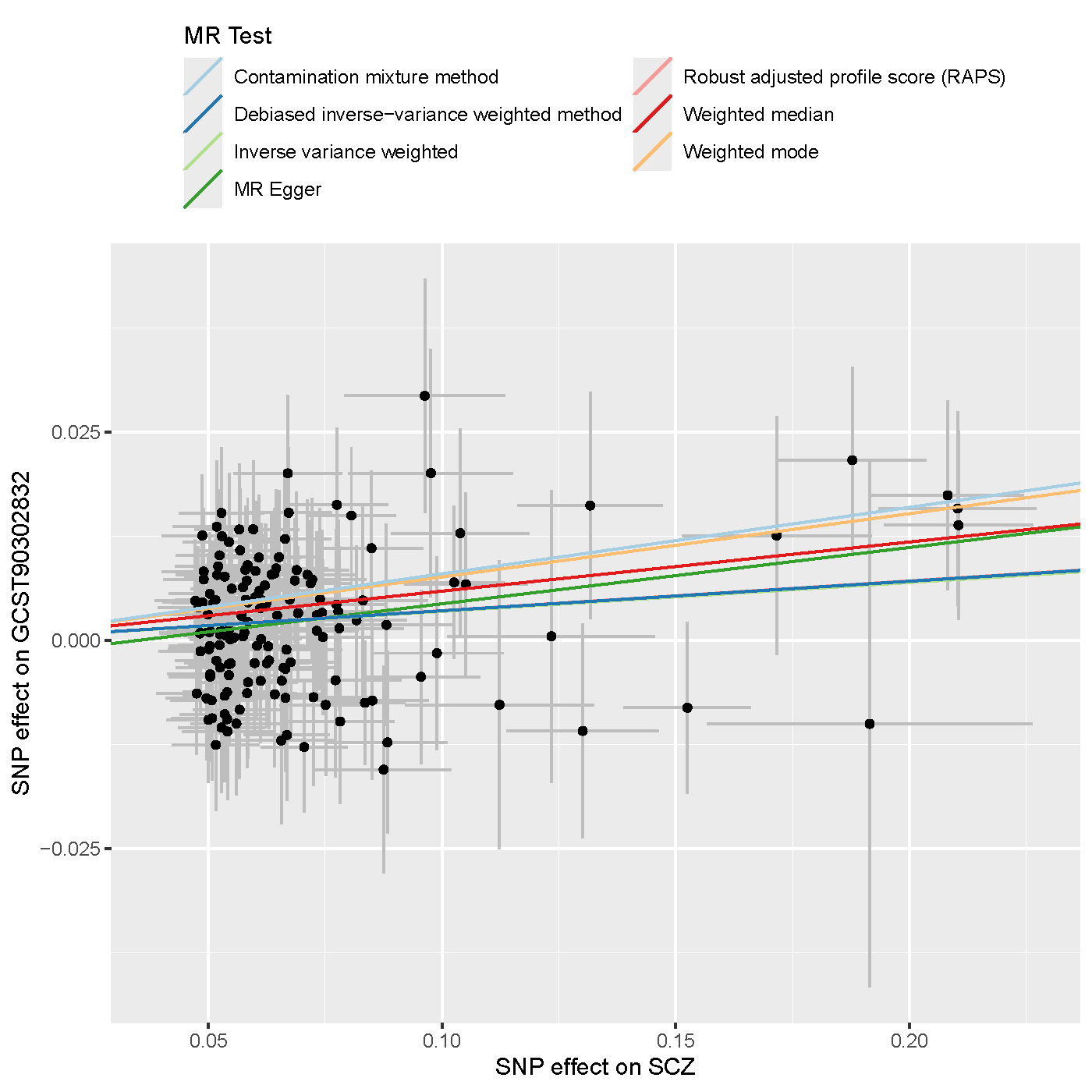

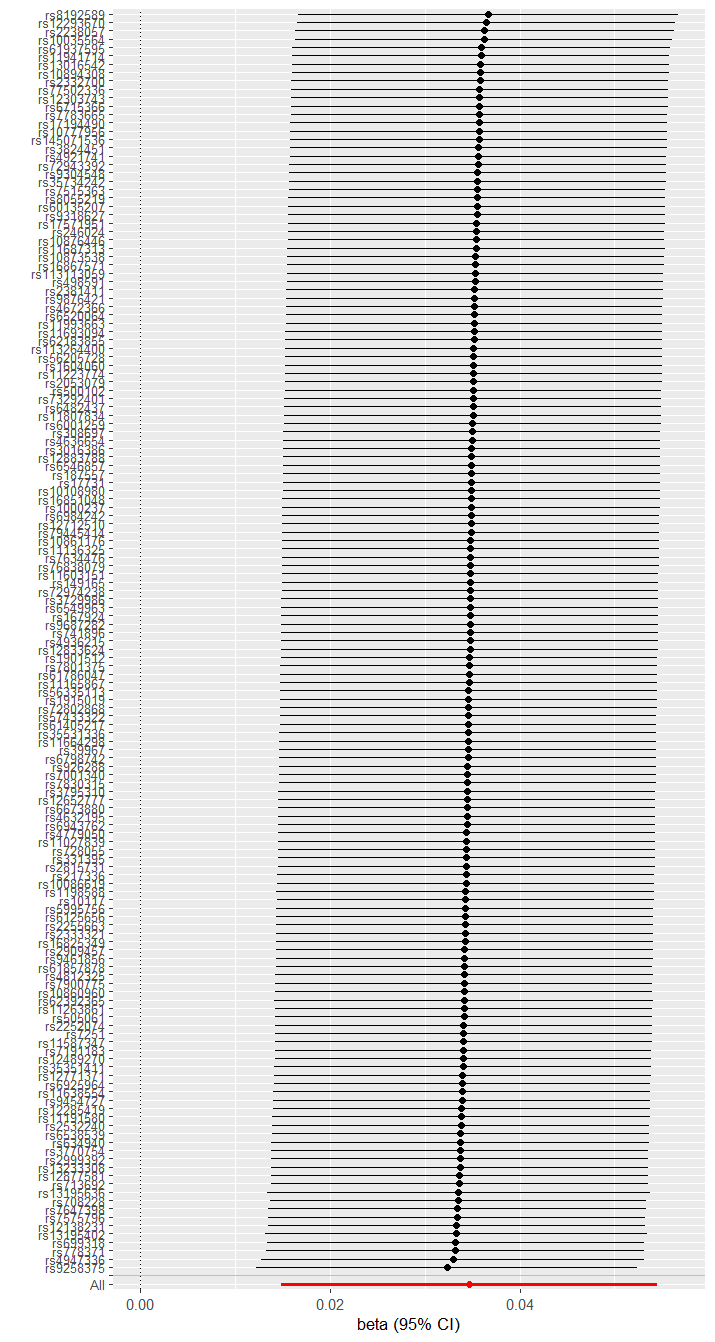

**Supplementary Figure 10 | SCZ & Right-hemisphere limbic network to amygdala white-matter structural connectivity (GCST90302832)**. **Left**: scatter plot displaying genetic associations with SCZ (x-axis) over genetic associations with GCST90302832 (y-axis) across seven different MR methods. **Right**: leave-one-out analysis plot of IVW results for SCZ-GCST90302832 exposure-outcome pair identified in reverse MR analyses, showing effect estimates for all SNPs and after excluding each SNP individually (y-axis), with beta (log odds ratio) and 95% confidence intervals (CI) on the x-axis.
